# Clinical Response to Azacitidine in Myelodysplastic Neoplasms is Associated with Distinct DNA Methylation Changes in Haematopoietic Stem and Progenitor cells *in vivo*

**DOI:** 10.1101/2024.07.19.24310679

**Authors:** Julie A.I. Thoms, Feng Yan, Henry R. Hampton, Sarah Davidson, Swapna Joshi, Jesslyn Saw, Chowdhury H. Sarowar, Xin Ying Lim, Andrea C. Nunez, Purvi M. Kakadia, Golam Sarower Bhuyan, Xiaoheng Zou, Mary Nguyen, Elaheh S. Ghodousi, Forrest C. Koch, Fatemeh Vafaee, Russell Pickford, Mark J. Raftery, Sally Hough, Griselda Buckland, Michelle Bailey, Yuvaraj Ghodke, Noorul Absar, Lachlin Vaughan, Leonardo Pasalic, Chun Y. Fong, Melita Kenealy, Devendra K. Hiwase, Rohanna I. Stoddart, Soma Mohammed, Linda Lee, Freda H. Passam, Stephen R. Larsen, Kevin J. Spring, Kristen K. Skarratt, Patricia Rebeiro, Peter Presgrave, William S. Stevenson, Silvia Ling, Campbell Tiley, Stephen J. Fuller, Fernando Roncolato, Anoop K. Enjeti, Dirk Hoenemann, Charlotte Lemech, Christopher J. Jolly, Stefan K. Bohlander, David J. Curtis, Jason W H Wong, Ashwin Unnikrishnan, Mark Hertzberg, Jake Olivier, Mark N. Polizzotto, John E. Pimanda

## Abstract

Hypomethylating agents are used as frontline therapy for myelodysplastic neoplasms (MDS), but clinical response is unpredictable. To determine whether response was associated with *in vivo* dynamics of DNA hypomethylation, we conducted a phase 2 trial for MDS using both injection and oral azacitidine (AZA). We established that global DNA methylation levels in peripheral blood and bone marrow mononuclear cells were comparable in AZA responders and non-responders during their course of treatment. However, there were distinct baseline and early drug induced differences in CpG methylation in haematopoietic stem and progenitor cells (HSPCs) in responders compared to non-responders that overlapped with regulatory regions of genes associated with tissue patterning, cell migration and myeloid differentiation. Following six cycles of therapy when clinical response typically manifests, differential hypomethylation in responder HSPCs pointed to marrow adaptation as a driver of enhanced haematopoiesis. Taken together, CpG methylation differences in HSPCs may explain variable response to AZA.

## Introduction

Myelodysplastic neoplasms (MDS) and chronic myelomonocytic leukemia (CMML) are clonal malignancies driven by accumulation of somatic mutations in haematopoietic stem cells (HSCs) which lead to impaired haematopoiesis and cytopenias along with increased likelihood of progressing to AML ^1–6^. High risk patients who are ineligible for haematopoietic stem cell transplant are treated with hypomethylating agents (HMAs), including azacitidine (AZA), which in a subset of patients leads to improved peripheral cell counts and delayed progression to AML ^7–10^. In the context of MDS/CMML, AZA has generally been administered by subcutaneous injection (Vidaza®, 75mg/m^2^/day), with a treatment cycle consisting of 7 days of injections followed by 21 days rest. More recently, an oral formulation (CC-486/Onureg®, 300mg/day for 14 days followed by 14 days rest) has been approved for maintenance therapy in AML patients who have achieved complete or incomplete remission following induction therapy and are not proceeding to a stem cell transplant ^11^. Clinical response to HMAs is generally not apparent until after 4-6 treatment cycles ^7^, and for patients with primary or secondary resistance to HMAs, treatment options are limited to enrolment in relevant clinical trials or supportive care.

Hypomethylating agents such as AZA and decitabine (DAC) are nucleoside analogues that, following intracellular conversion processes ^12,13^, are incorporated into DNA and, in the case of AZA, into RNA ^14^. HMA incorporation into DNA requires that target cells are cycling ^15^, and we have previously reported a correlation between cell cycle parameters at diagnosis and subsequent patient response ^16^. Once incorporated into DNA, HMAs trap the maintenance DNA methyltransferase DNMT1 leading to formation of DNA-protein adducts, degradation and cellular depletion of DNMT1, and subsequent global hypomethylation ^17–20^. Multiple stem-cell-intrinsic effects of HMAs observed in patients have been proposed to mediate the clinical effects of HMAs including epigenetic reactivation of tumour suppressor genes ^21,22^, activation of silenced retroviral elements and subsequent viral mimicry response ^23–25^, DNA-damage response and related cytotoxicity ^17,18,26^, induction of apoptosis ^27–29^, and RNA-dependent mechanisms ^30,13,31,32^. However, HMA-mediated cytotoxicity is not essential for clinical response ^33,34^, and there is emerging evidence that response to HMAs (in the context of MDS/CMML) does not require complete eradication of mutated clones; rather that improved peripheral blood counts are driven by increased output from mutated stem cells ^35,36,16,37^. Overall, no straightforward relationship has been demonstrated between stem-cell-intrinsic effects of HMA treatment and patient outcome ^21,38,39,16,40,41^, rendering prediction of response status and development of rationally designed combination therapies an ongoing challenge.

In particular, the association between HMA incorporation and DNA hypomethylation, and the clinical efficacy of these drugs is unclear. In a small retrospective study, HMA incorporation in a single initial treatment cycle tracked with global demethylation, with more variable uptake and hypomethylation observed in patients who did not subsequently respond ^41^. However, HMA incorporation and DNA hypomethylation kinetics have not previously been studied over the time span where clinical response becomes evident. Furthermore, these parameters have not been assessed in parallel with cell cycle characteristics or changes in clonal composition during HMA treatment. To address these relationships, we prospectively collected bone marrow (BM) and peripheral blood (PB) as part of a phase II clinical trial (NCT03493646) designed to evaluate *in vivo* AZA incorporation in mononuclear cells following treatment with Vidaza (injection AZA) or CC-486 (oral AZA). The primary objective of the trial was to assess drug incorporation into DNA and global changes in DNA methylation using an LC-MS/MS based method ^41^, while secondary objectives included observation of cell cycle changes in haematopoietic stem and progenitor cells (HSPCs), tracking of clonal variants, and assessment of any relationship between these measures and clinical response.

## Results

### Patient cohort and clinical outcomes

Patients newly diagnosed with high risk (HR) MDS, CMML, or low blast AML, and ineligible for allogeneic stem cell transplant or intensive chemotherapy (Table S1), were enrolled to an open label phase 2 multicentre investigator-initiated trial (NCT03493646) and administered six cycles of injection AZA followed by six cycles of oral AZA (Figure 1A). Clinical response was assessed following cycle 6 (C6D28) and cycle 12 (C12D28) using IWG2006 criteria ^42^. Forty patients commenced treatment. Of the 24 who completed 6 cycles of injection AZA, there were 16 responders (seven complete remission (CR), three marrow complete response (mCR), and six hematological improvement (HI)) and eight non-responders (five stable disease (SD), three progressive disease (PD)/failures) (Figure 1B, Figure 1C, Table S2). All participants were retrospectively assessed using IWG2023 criteria ^43^; of these, one patient (P10) changed from non-responder to responder at C12D28 (Table S3).

**Figure 1:**
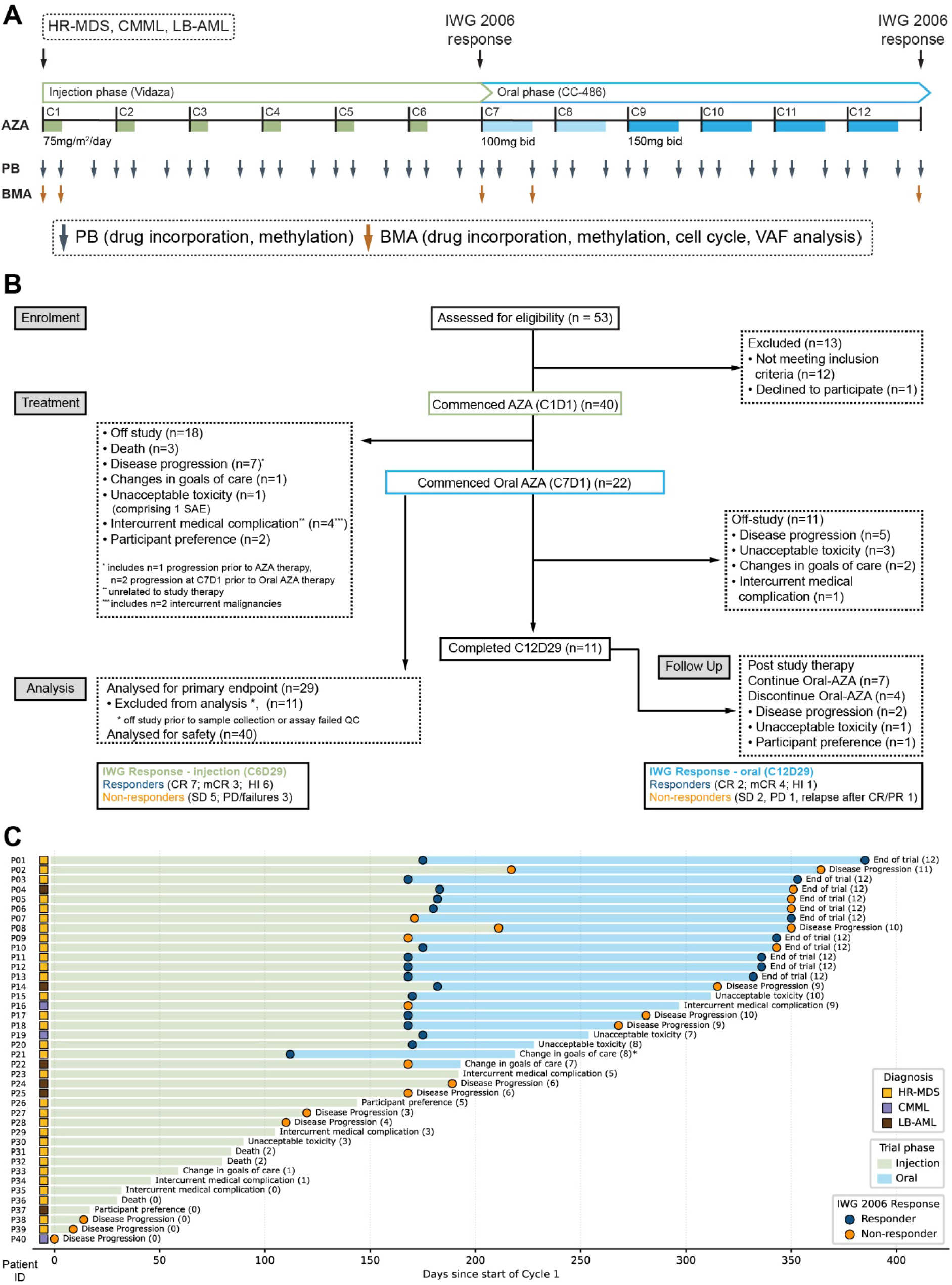
Trial design and patient outcomes. **(A)** Trial design showing dosing schedule, specimen collection, and IWG assessment timepoints for injected and oral phases. **(B)** Consort diagram summarising outcomes for all participants who were assessed for eligibility. **(C)** Swimmer plots showing participant response and outcome over time. Colored squares indicate diagnosis for each patient (yellow – high risk (HR) MDS, purple – CMML, brown – low blast (LB) AML). Circles show response at IWG assessment or progression timepoints (blue – responder, orange – non-responder). The off-study reason for each patient is as indicated, the number in brackets indicates the number of treatment cycles completed, * denotes a single patient (P21) who was accelerated to the oral phase following 4 injection cycles.

Twenty-two patients entered the oral AZA phase and completed median 2 cycles (range 0-6); a total of 100 cycles of oral AZA were delivered. Reasons for discontinuation during the oral AZA phase (n=11) included disease progression (five patients, 46%), unacceptable toxicity (three patients, 27%; one haematological, two gastrointestinal), changes in goals of care (two patients, 18%), and intercurrent medical complication (one patient, 9%). The most common adverse events during oral AZA therapy were haematological (Table S4, Table S5): 23 cycles in 13 patients were complicated by grade 4 neutropenia, one cycle in one patient by febrile neutropenia, and 14 cycles in 10 patients by grade 4 thrombocytopenia. Other adverse events were less common, with 13 cycles in nine patients complicated by grade 3 gastrointestinal toxicity (diarrhoea; five events in three participants, nausea; four events in three participants, rectal haemorrhage; two events in one participant, one event each for vomiting and abdominal pain; no grade 4 adverse events were observed).

11/22 patients who commenced oral AZA reached the second response assessment (C12D28) comprising seven responders (two CR, four mCR, one HI) and four non-responders (two SD, one PD, one relapse after CR/PR). The seven responders at C12D28 included five responders and two non-responders at C6D28 (i.e. two patients who showed a delayed clinical response), while the four non-responders at C12D28 had previously shown response at C6D28 (Figure 1C). Most patients completing cycle 12 elected to continue to receive treatment with oral AZA (64%, 7/11).

### Cell cycle parameters at diagnosis and over the course of treatment do not correlate directly with clinical outcome

Previous studies suggested that patients who respond to AZA have a higher proportion of actively cycling (S/G_2_/M phase) CD34^+^CD38^hi^ hematopoietic progenitor cells (HPC) at baseline compared to patients who do not have a clinical response ^16^. We measured cell cycle parameters in bone marrow CD34^+^CD38^lo^ HSC and HPC across the course of AZA treatment (Figure 2A). In HSCs, there were few actively cycling cells at baseline regardless of subsequent clinical response (median % in cell cycle phase, [interquartile range]; Responders (R): [0.5, (0.1-1.2)]; Non-responders (NR): [0.2, (0.1-0.5)]), and the overall proportion of cells in G_0_ or G_1_ were similar between response groups (Figure 2Bi,ii,iii, *left panels*; G_0_ - R: [83.2, (68.7-90.7)]; NR: [57.7, (48.6-91.4)]; G_1_ - R: [16.6, (6.3-25.6)]; NR: [35.3, (7.4-44.6)]). Following six cycles of injection AZA the proportion of HSCs in S/G_2_/M was relatively stable (R: [0.5, (0.0-0.9)]; NR: [1.2, (0.6-1.4)]). There was an overall decrease in the proportion of quiescent G_0_ cells (R: [53.8, (47.2-60.8)]; NR: [56.8, (44.6-61.2)]) and concomitant increase in the proportion of cells in G_1_ (R: [42.6, (37.5-48.5)]; NR: [40.9, (35.0-53.4)]), but this difference reached significance only in responders (Figure 2Bi,ii,iii, *right panels*). In HPCs, there was a trend for responder patients to have an increased proportion of cells in S/G2/M at baseline (R: [5.0, (2.9-5.7)]; NR: [3.2, (2.2-6.0)]), although this did not reach statistical significance. Similar to HSCs, the overall proportion of G_0_ and G_1_ cells were similar between response groups at baseline (Figure 2Ci,ii,iii; G_0_ - R: [59.9, (55.8-69.8)]; NR: [48.4, (29.8-71.2)]; G_1_ - R: [32.9, (25.0-38.6)]; NR: [48.5, (24.0-61.5)]. After six treatment cycles, some patients had shifts in the proportion of HPCs in S/G_2_/M, but there was no consistent change either across the entire cohort, or within response groups (R: [7.2, (4.1-9.9)]; NR: [7.3, (5.7-9.2)]). However, similar to HSCs there was an overall decrease in the proportion of quiescent cells (G_0_ - R: [23.5, (17.3-38.7)]; NR: [35.2, (24.2-40.4)]) and an increase in the proportion of G_1_ cells (R: [69.4, (56.4-72.8)]; NR: [57.5, (54.4-66.1)]) both of which were significant only in responders (Figure 2Ci,ii,iii, *right panels*).

**Figure 2:**
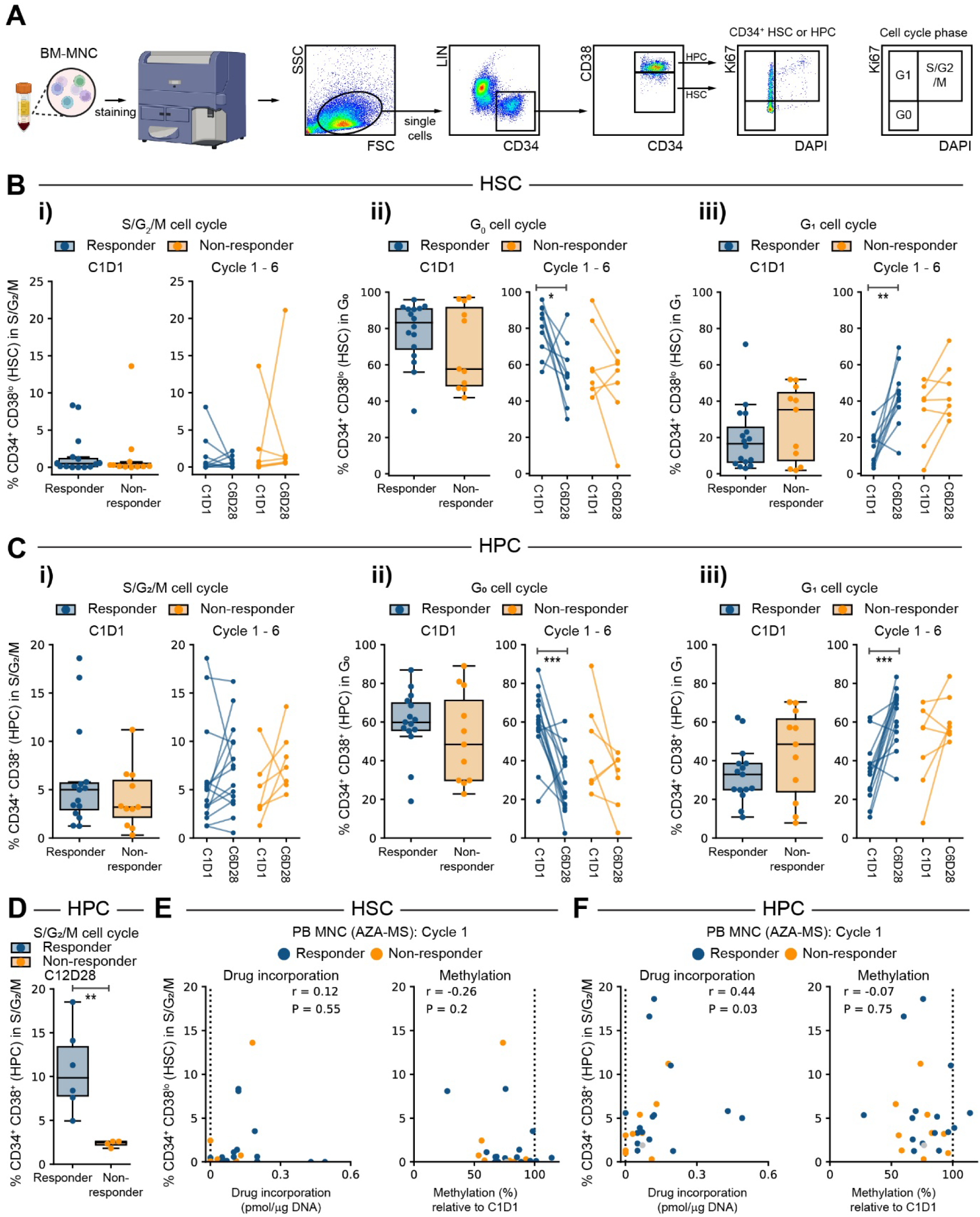
Cell cycle status at diagnosis and across treatment cycles. **(A)** Schematic showing analysis workflow for determining cell cycle parameters in CD34^+^ CD38^lo^ stem (HSC) and CD34^+^ CD38^hi^ progenitor (HPC) cells. Schematic partially created with BioRender.com **(B)** Percentage of HSCs in **(i)** S/G_2_/M, **(ii)** G_0_, **(iii)** G_1_ cell cycle phases. **(i-iii)** *Left:* percentage of cells in specified cell cycle phase at diagnosis (C1D1), coloured by clinical outcome at the end of the injection phase. *Right:* Percentage of cells in specified cell cycle phase at diagnosis (C1D1) and at the end (C6D28) of the injection phase. Lines indicate paired samples from a single patient, only patients with data at C1 and C7 are shown. *** *P* < 0.001, ** *P* < 0.01, * *P* < 0.05 unpaired (*left*) or paired (*right*) t-test. **(C)** Percentage of HPCs in **(i)** S/G_2_/M, **(ii)** G_0_, **(iii)** G_1_ cell cycle phases. **(i-iii)** *Left:* percentage of cells in specified cell cycle phase at diagnosis (C1D1), coloured by clinical outcome at the end of the injection phase. *Right:* Percentage of cells in specified cell cycle phase at diagnosis (C1D1) and at the end (C6D28) of the injection phase. Lines indicate paired samples from a single patient, only patients with data at C1 and C7 are shown. *** *P* < 0.001, ** *P* < 0.01, * *P* < 0.05 unpaired (*left*) or paired (*right*) t-test. **(D)** Percentage of HPCs in S/G_2_/M following 12 treatment cycles, coloured by clinical outcome at C12D28. *** *P* < 0.001, ** *P* < 0.01, * *P* < 0.05 unpaired t-test. **(E-F)** Relationship between cell cycle status and drug incorporation and DNA demethylation during cycle 1 in **(E)** HSCs and **(F)** HPCs. **(E-F)** Percentage of cells actively cycling (S/G_2_/M phase) at C1D1 compared to maximum drug incorporation (*left*) and minimum DNA methylation (*right*) in peripheral blood (PB) during the same cycle. Dashed lines indicate baseline values. Gray dots indicate patients for whom response data is unavailable. r = spearman correlation coefficient.

At the start of the oral phase (C7D1), HSC cell cycle parameters were similar between responders and non-responders (Figure S1Ai,ii,iii; S/G_2_/M - R: [0.6, (0.3-1.1)]; NR: [0.7, (0.3-1.3)]; G_0_ - R: [47.9, (38.5-58.5)]; NR: [53.8, (47.3-60.6)]; G_1_ - R: [43.7, (39.4-59.4)]; NR: [43.2, (37.1-51.5)]). For some patients, the quantity of HSCs was too low (<50 phenotypic HSCs) to reliably measure percentage of cells in each cell cycle phase, limiting analysis of paired samples. Cell cycle parameters in HPCs at the start of the oral phase were also similar between response groups (Figure S1Bi,ii,iii; S/G_2_/M - R: [5.4, (4.3-7.3)]; NR: [8.2, (4.2-9.9)]; G_0_ - R: [21.6, (17.0-28.6)]; NR: [39.5, (17.4-41.1)], G_1_ - R: [72.2, (67.1-73.6)]; NR: [57.8, (52.1-72.6)]). There was a trend for responder patients to have an increase, and non-responder patients have a decrease in the proportion of cells in S/G_2_/M over the oral phase; strikingly, by the end of treatment, HPCs in non-responder patients had essentially stopped cycling (Figure 2D, Figure S1C; R: [9.9, (7.8-13.4)]; NR: [2.5, (2.2-2.6)]).

Cell cycle kinetics might directly influence the amount of DAC incorporated into DNA, and therefore the extent of DNA demethylation. We compared cell cycle parameters with DAC incorporation and global DNA demethylation in circulating cells during cycle 1 in HSCs (Figure 2E) and HPCs (Figure 2F). Immediately following a single treatment cycle DAC incorporation and demethylation were observed in essentially all patients, and the degree of DAC incorporation was correlated to the proportion of HPCs in S/G2/M (Figure 2F; Spearman r = 0.44, *P* = 0.03). During cycle 7 (oral phase) DAC incorporation and demethylation were again observed in essentially all patients, however neither correlated with cell cycle parameters at this time point (Figure S1D, Figure S1E). Overall, we found that most patients had fewer quiescent cells following six cycles of AZA treatment, and at the end of cycle 12, HPCs in responder patients continued to cycle, while HPCs in non-responders had exited the cell cycle. However, cell cycle parameters did not directly correlate with clinical outcome.

### DAC incorporation and global DNA demethylation in peripheral blood and bone marrow are not correlated with clinical response

AZA undergoes intracellular modification prior to being incorporated into newly synthesised DNA, where it traps the maintenance methyltransferase DNMT1 leading to DNMT1 degradation and DNA hypomethylation ^19^ (Figure 3A). We directly measured drug incorporation into DNA and relative global DNA methylation in PB mononuclear cells (MNCs) at 1-2 weekly intervals during both injection and oral treatment phases (Figure 1A). During the injection AZA phase, drug incorporation was higher in responders [0.093 (95% CI 0.066-0.120)] compared to non-responders [0.045 (95% CI 0.011 – 0.079)] pmol DAC/µg DNA; p = 0.032 (Figure 3B). Analysis of drug incorporation at day 1 (D1), day 8 (D8) and day 22 (D22) revealed that incorporation differences were primarily driven by D8 measurements, i.e. immediately following 7 days of AZA treatment (Figure 3B). However, overall differences in DNA methylation relative to baseline (100%) were comparable between responders [85.41% (95% CI 77.40-93.43)] and non-responders [86.85% (95% CI 76.69 – 97.01] (p= 0.83), with peak demethylation observed at D8 in both treatment groups (Figure 3C, Figure S2A-D).

**Figure 3:**
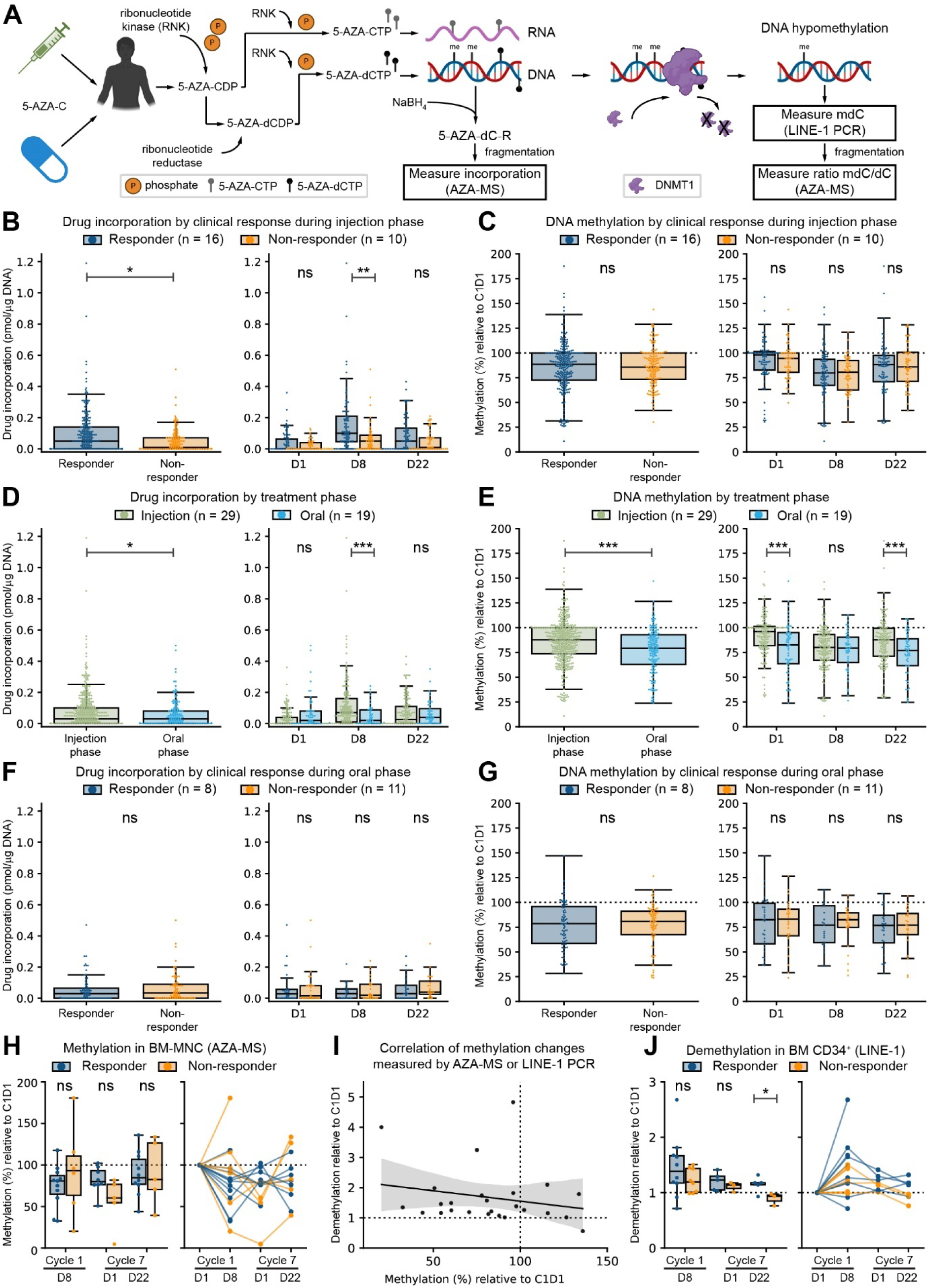
DAC incorporation and relative DNA methylation across treatment phases. **(A)** Schematic showing administration of azacitidine (5-AZA-C), *in vivo* modification, and incorporation into DNA leading to trapped and degraded DNMT1 and DNA hypomethylation. Drug incorporation into DNA is directly assessed by measuring NaBH_4_-stabilised and fragmented DNA (dihydro 5-AZA-dC ribonucleotide (R)) by mass spectrometry. Changes in DNA methylation were assessed either by mass spectrometry-based comparison of mdC to dC ratio pre- and post-treatment or by measuring demethylation of LINE-1 promoter elements as a proxy for global demethylation. Schematic partially created with BioRender.com **(B-C)** Drug incorporation and DNA methylation (relative to baseline) during the injection phase in patients who either progressed or completed the treatment phase and underwent IWG disease assessment to determine response status (n = 26 [Responder (R): 16, Non-responder (NR): 10]). **(B)** Drug incorporation in responders and non-responders at all sample points (*left*) or separated by cycle day (*right*). All statistical comparisons for panels B-G were performed using a linear mixed model approach, *P* values are as indicated (*** *P* < 0.001, ** *P* < 0.01, * *P* < 0.05, ns *P* ≥ 0.05). **(C)** Relative DNA methylation at all sample points (*left*) or separated by cycle day (*right*). **(D-E)** Drug incorporation and DNA methylation during the injection and oral phases of the study (n = 29 patients during injection phase, n = 19 patients during oral phase); data analysis for the oral phase was restricted to cycles 9 and later to minimise the likelihood of detecting changes carried over from the injection phases. **(D)** DAC incorporation during the injection and oral phase at all sample points (*left*) or separated by cycle day (*right*). **(E)** Relative DNA methylation at all sample points (*left*) or separated by cycle day (*right*). **(F-G)** Drug incorporation and DNA methylation during the oral phase of the study: data analysis was restricted to cycles 9 and later to minimise the likelihood of detecting changes carried over from the injection phases (n = 19 patients, R = 8, NR = 11). Response status is based on IWG assessment at end of cycle 12 when available (n = 11); patients who progressed during the oral phase or soon after stopping treatment were assessed as non-responders (n = 6), and remaining patients were assigned response as per end of cycle 6 IWG assessment (n = 2). **(F)** DAC incorporation in responders and non-responders at all sample points (*left*) or separated by cycle day (*right*). **(G)** Relative DNA methylation at all sample points (*left*) or separated by cycle day (*right*). **(H)** Relative DNA methylation in bone marrow mononuclear cells at C1D8, C7D1, and C7D22 compared to pre-treatment. *Left:* aggregate data at each timepoint. *Right:* Plot showing longitudinal methylation changes in individual patients. Statistical comparisons for panels H and J were performed using the ttest_ind function from scipy.stats, *P* values are as indicated * *P* < 0.05, ns *P* ≥ 0.05) **(I)** Comparison of methylation changes measured by mass spectrometry or LINE-1 PCR assay in bone marrow mononuclear cells. Individual samples are plotted along with the linear regression line with 95% confidence interval. **(J)** Relative DNA demethylation in bone marrow CD34^+^ cells at C1D8, C7D1, and C7D22 compared to pre-treatment. *Left:* aggregate data at each timepoint. *Right:* Plot showing longitudinal methylation changes in individual patients.

Nineteen patients completed at least one cycle of oral AZA at 150 mg bid (i.e., completed C9) and were compared to all patients with injection AZA data (n = 29) for assessment of the primary outcome. Drug incorporation was higher during the injection phase (Figure 3D: injection AZA; 0.070 (95% CI 0.052 – 0.089) vs. oral AZA; 0.047 (95% CI 0.023 – 0.070) pmol DAC/µg DNA, p = 0.02), again primarily driven by D8 measurements (Figure 3D). However, there was lower overall DNA methylation during the oral phase (Figure 3E: injection AZA; 86.42% (95% CI 81.06 – 91.78) vs. oral AZA; 77.64% (95% CI 71.67 – 83.61); p <0.0001). Methylation differences were significant at both D1 and D22, but not at D8 (Figure 3E). Thus, although we observed increased drug incorporation with injection AZA, there was greater and more sustained demethylation in response to oral administration of AZA.

During the oral phase, drug incorporation was comparable between responders and non-responders (Figure 3F: responders; 0.059 (95% CI 0.012 – 0.106) vs. non-responders; AZA 0.073 (95% CI 0.031 – 0.115) pmol DAC/µg DNA, p = 0.66). Similar to injection AZA, we did not observe any methylation differences between the response groups (Figure 3G: responders; 79.61% (95% CI 62.35 – 96.87) vs. non-responders; 79.23% (95% CI 64.31 – 94.15); p = 0.97).

Although DNA demethylation is a likely mediator of the therapeutic effects of AZA, in our cohort essentially every patient had reduced DNA methylation in peripheral blood in response to treatment, and there was no direct relationship between drug incorporation, level of global DNA demethylation, and clinical outcome (Figure 3C, Figure 3G). One possibility is that the clinically relevant demethylation events occur in specific cells within the bone marrow and may not be read out in circulating cells. We first measured global DNA demethylation in BM MNCs before and after AZA treatment in cycle 1 and cycle 7. Similar to PB, demethylation was apparent in BM MNC from the majority of patients and was not significantly different between responders and non-responders (Figure 3H).

MDS/CMML are diseases of stem cells, and clinical response in this cohort is measured in improved circulating blood counts likely driven by improved output from CD34^+^ HSPCs ^36^. HSPCs are rare, and not amenable to AZA-MS analysis. However, relative methylation of LINE-1 promoters as a proxy measure of global methylation changes can be assessed in small cell numbers using a PCR-based assay ^44^ (Figure 3I, Figure S2E). Comparing CD34^+^ cells from responders and non-responders, there was a trend for greater demethylation in responders which reached statistical significance at C7D22 (Figure 3J). Taken together, our data indicated that most patients treated with AZA undergo drug incorporation and global DNA demethylation, and that the level of global demethylation in CD34^+^ HSPCs may be related to clinical response.

### Pre-treatment DNA methylation at specific CpG sites correlates with clinical response

To further investigate the relationship between DNA methylation changes and clinical outcome we performed reduced representation bisulfite sequencing (RRBS) on longitudinal samples of BM CD34^+^ cells prior to, and immediately following, AZA treatment during cycle 1 and cycle 7 (Figure 4A). At baseline (C1D1), the number of CpGs detected with more than 10 reads (Figure S3A; range 2.4 - 3.6 million) and total reads in detected CpGs (Figure S3A; range 38.6 - 114.9 million) were similar between samples, and hierarchical clustering of all samples showed highest similarity within samples derived from the same patient (Figure S3B). Patients with MDS/CMML/AML generally have global hypermethylation ^45^. However, at baseline (C1D1), we observed relative hypomethylation in CD34^+^ cells of patients who responded to AZA compared to patients who did not respond (Figure 4B, Figure S3C), suggesting that differences in DNA methylation at diagnosis might influence patient outcomes. 23950 CpGs were hypomethylated in responders compared to non-responders and were predominantly located in CpG islands and promoter regions, while 3004 CpGs were hypermethylated in responders and were more frequently located in distal regions (Figure 4C). Since cytosine methylation is often concordant in proximity, we combined differentially methylated cytosines (DMCs) to regions (DMRs) prior to clustering baseline data. Patients clustered by response group, with the average methylation percentage of DMRs remaining consistent within response groups, indicating shared regions of differential methylation between individuals (Figure 4D). We then linked DMCs to target genes using HiChIP data generated from healthy HSPCs ^46^ and used pathway analysis to predict functional consequences of differential methylation between responders and non-responders. Genes associated with hypomethylated DMCs in responders included HOXA and HOXB cluster genes, GATA2, and SNAI1 (Figure 4E). The top 10 gene ontology analysis hits included pattern specification processes and regulation of epithelial cell migration (Figure S3Di); specific gene hits and their inclusion in selected pathways are shown (Figure 4E).

**Figure 4:**
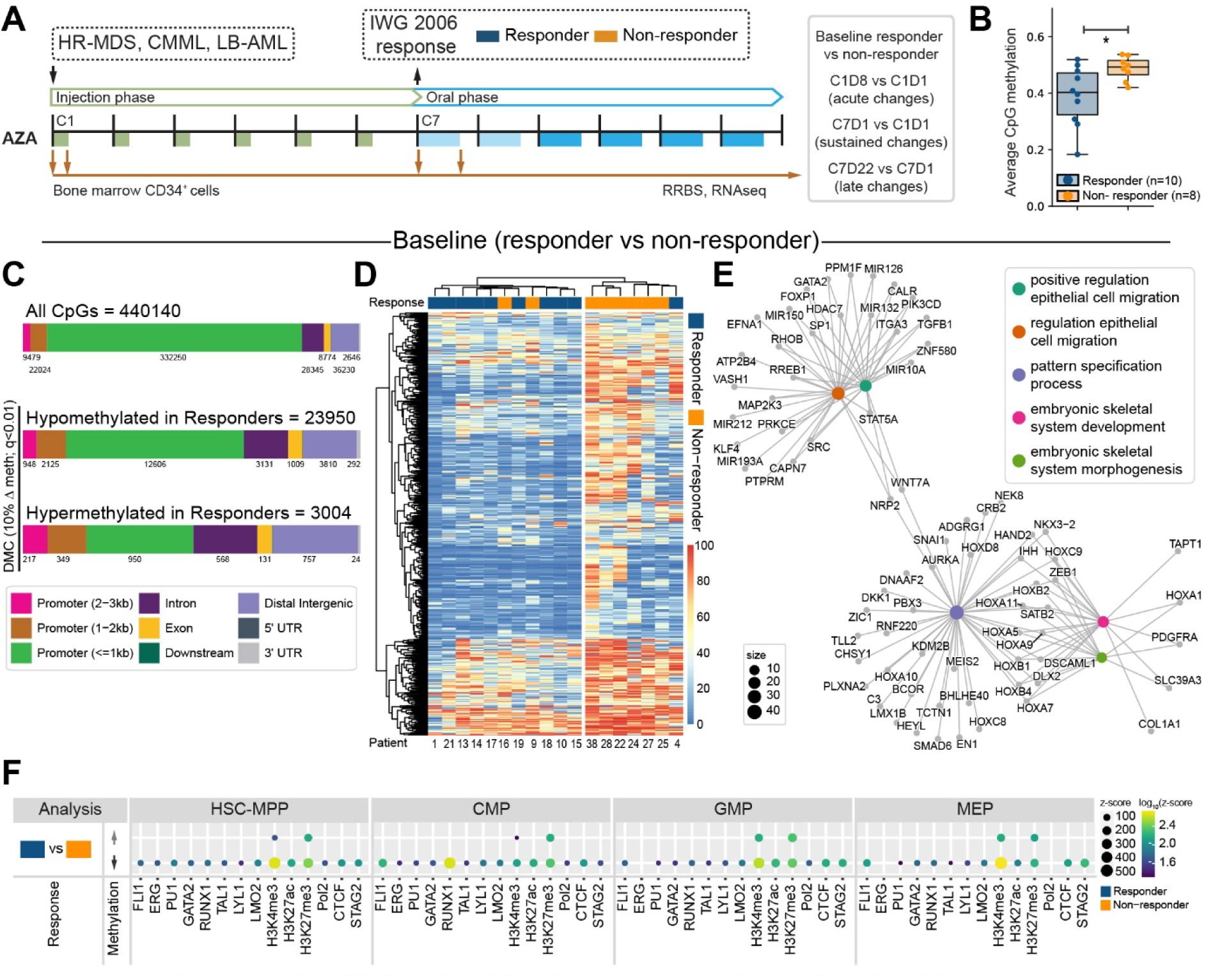
Baseline differences in site-specific methylation correlate with clinical response to AZA. **(A)** Schematic showing sample collection timepoints for reduced representation bisulphite sequencing (RRBS). **(B)** Average methylation at CpG sites at baseline (C1D1). Statistical comparison is unpaired t-test, * indicates *P* < 0.05. **(C-F**) Comparison of responder (n = 10) and non-responder (n = 8) patients prior to commencing AZA therapy. **(C)** Upper bar shows genomic distribution of 440140 CpG sites with data in all samples at C1D1 and C1D8. Lower bars show genomic distribution of CpGs which were hypomethylated (n = 23950) or hypermethylated (n = 3004) in responders compared to non-responders. **(D)** Clustered heatmap showing differentially methylated regions across all patients, with clinical response as indicated. **(E)** Network diagram showing enriched pathways for genes mapping to CpGs which are hypomethylated in responders at baseline. Network diagrams were created using clusterProfiler. CpGs were annotated to genes using HiChIP data from healthy human HSPC subsets ^46^. **(F)** Enrichment of differentially methylated CpGs at genomic regions with specific histone marks (H3K27Ac, H3K3me3, H3K27me3) or bound by key transcription factors and genome organisers (FLI1, ERG, PU.1, GATA2, RUNX1, TAL1, LYL1, LMO2, Pol2, CTCF, STAG2) in healthy human HSPC subsets. Plot shows CpG regions hypermethylated (*upper*) or hypomethylated (*lower*) in responders compared to non-responders.

Baseline differences in CpG methylation suggests that CD34^+^ HSPCs might be epigenetically primed for AZA response in a subset of patients. To better understand how hypomethylated DMCs in responders might influence the global epigenetic environment, we overlapped baseline DMCs with global transcription factor (TF) and histone ChIPseq data from healthy HSPC subsets (HSC-multipotent progenitors (MPP), common myeloid progenitor (CMP), granulocyte-monocyte progenitor (GMP), megakaryocyte-erythroid progenitor (MEP)) ^47^. There was striking overlap between TF/histone binding and DMCs hypomethylated in responders, but only at select histone marks for DMCs hypermethylated in responders (Figure 4F). Both hypo- and hyper-methylated DMCs showed overlap with H3K4me3 (promoter mark) and H3K27me3 (repressive mark) which together mark bivalent regions primed for epigenetic plasticity ^48,49^. CTCF is involved in defining chromatin boundaries, and along with STAG2 belongs to the cohesin complex which facilitates looping of promoters to distal regulatory regions ^50,51^. We observed enriched overlap with CTCF and STAG2 binding sites only at DMCs that were hypomethylated in responders (Figure 4F). Finally, overlap with TF binding sites was most prominent at FLI1-bound regions in CMPs and MEPs, and RUNX1-bound regions in CMPs. In CMPs and MEPs both FLI1 and RUNX1 bind regulatory regions of lineage specific genes that are subsequently expressed in mature myeloid or erythroid cells ^46^ which are the specific cell populations that contribute to clinical response. Overall, we observe significant baseline CpG hypomethylation in patients who go on to respond to AZA, with the hypomethylated regions overlapping genes critical for blood development, and regulatory sites associated with epigenetic plasticity and lineage-specific gene regulation.

### Longitudinal assessment of DNA methylation shows dynamic changes at specific CpG sites correlate with clinical response

We next looked at acute methylation changes immediately following the initial seven days of AZA treatment. Extensive hypomethylation was evident across all patients, with more than twice as many CpGs hypomethylated in responders compared to non-responders (Figure 5A; 23060 vs 9580). Hypermethylation compared to baseline was also observed at a very small number of CpGs (Figure 5A; 79 vs 40). While baseline methylation differences were mostly promoter-proximal, hypomethylation following AZA treatment was mostly at distal regions or within introns, regions which potentially correspond to enhancers (Figure 5A). We next compared overlap of specific DMCs between responders and non-responders (Figure 5B). Notably, 7572 CpGs were hypomethylated in both responders and non-responders; these comprised the majority of DMCs in non-responders. Gene ontology analysis of genes associated with the shared hypomethylated CpGs revealed a single enriched pathway (Figure S3Dii; immune response regulating signalling pathway). A further 15488 CpGs were uniquely hypomethylated in responders, while 2005 CpGs were uniquely hypomethylated in non-responders. Importantly, genes associated with CpGs uniquely hypomethylated in responders were enriched for pathways relating to myeloid cell and osteoclast differentiation (Figure S3Diii, Figure 5C) suggesting clinical response to AZA is at least partly due to epigenetic reprogramming at gene loci required for HSPCs to differentiate and produce circulating progeny. Overlap of hypomethylated CpGs in responders at C1D8 with global transcription factor (TF) and histone ChIPseq data from healthy HSPC subsets revealed enrichment at regulatory regions, particularly at H3K27ac sites (active enhancer) in GMPs and MEPs, again consistent with improved differentiation capacity (Figure 5D).

**Figure 5:**
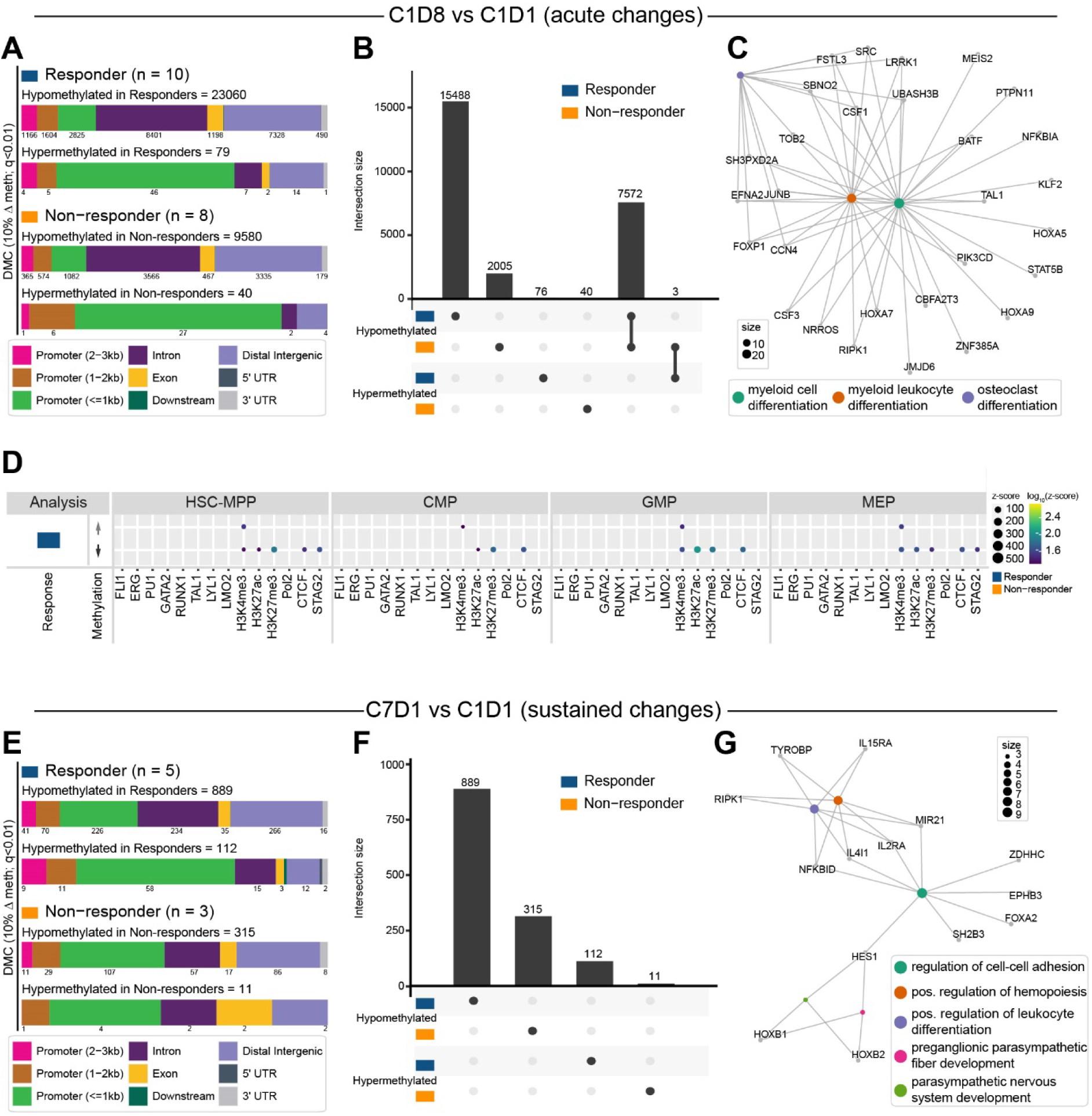
Acute and sustained changes in site-specific methylation following AZA treatment. **(A-D)** Acute methylation changes at 440140 CpG sites detected in all samples at C1D1 and C1D8 following the first cycle of AZA treatment. **(A)** Genomic distribution of CpG sites differentially methylated at C1D8 compared to baseline C1D1 in responders and non-responders. **(B)** Upset plot showing overlap of differentially methylated CpG sites between comparison groups. **(C)** Network diagram showing enriched pathways for genes mapping to CpGs which are uniquely hypomethylated in responders at C1D8 compared to C1D1 (n = 15488 CpGs). Network diagrams were created using clusterProfiler. CpGs were annotated to genes using HiChIP data from healthy human HSPC subsets ^46^. **(D)** Enrichment of differentially methylated CpGs at genomic regions with specific histone marks (H3K27Ac, H3K3me3, H3K27me3) or bound by key transcription factors and genome organisers (FLI1, ERG, PU.1, GATA2, RUNX1, TAL1, LYL1, LMO2, Pol2, CTCF, STAG2) in healthy human HSPC subsets. Plot shows CpG regions hypermethylated (*upper*) or hypomethylated (*lower*) at C1D8 compared to C1D1 in responders. **(E-G)** Sustained changes in site-specific methylation. **(E)** Genomic distribution of CpG sites differentially methylated at C7D1 compared to baseline in responders and non-responders. **(F)** Upset plot showing overlap of differentially methylated CpG sites between comparison groups. **(G)** Network diagram showing enriched pathways for genes mapping to CpGs which are hypomethylated in responders at C7D1 compared to C1D1 (n = 889 CpGs). Network diagrams were created using clusterProfiler. CpGs were annotated to genes using HiChIP data from healthy human HSPC subsets ^46^.

Clinical response to AZA is generally not evident for several months following commencement of treatment ^7^, and the substantial changes in CpG methylation that occur immediately following the initial treatment cycle may not persist over the long term. To evaluate sustained changes in CpG methylation we compared CpG methylation at C7D1 (after 6 AZA treatment cycles, but 3 weeks after the last AZA dose) to baseline methylation. Hypomethylation relative to baseline was observed in all patients, again skewed to distal regulatory regions, with approximately 3 times as many sites hypomethylated in responders compared to non-responders (Figure 5E; 889 vs 315). However, the overall number of hypomethylated sites was significantly reduced compared to C1D8, and there was no overlap in hypomethylated sites between response groups (Figure 5F). Gene ontology analysis of genes associated with hypomethylated CpGs in responders revealed enrichment in multiple pathways including regulation of cell-cell adhesion and positive regulation of hemopoiesis; these pathways may reflect a marrow environment that is more supportive of blood production (Figure S3Div, Figure 5G).

Finally, to assess late AZA-responses in patients following long term AZA treatment we compared CpG methylation at C7D22 to C7D1 (Figure S3E). Surprisingly, we observed mostly hypermethylation in all patients, albeit at a very small number of genes (Figure S3E; responders - 106, non-responders - 83). Only 18 CpGs were hypomethylated in responders, and there was minimal overlap between treatment groups except for a single shared hypermethylated CpG (Figure S3F). Genes associated with the 18 CpGs hypomethylated in responders were enriched in pathways including regulation of signal transduction by p53 class mediator and regulation of mitotic cell cycle phase transition (Figure S3Dv, Figure S3G).

Overall, we observed that patients with clinical response to AZA have lower CpG methylation at baseline and undergo further CpG hypomethylation at regions associated with genes involved in myeloid cell differentiation. Furthermore, changes in CpG methylation that persist following 6 cycles of AZA treatment are also enriched for genes involved in haematopoiesis and leukocyte differentiation.

### Variant alleles persist over the course of AZA treatment irrespective of clinical response

Improved peripheral blood counts, which are the hallmark of clinical response to MDS/CMML, arise via improved blood production originating from CD34^+^ HSPCs. An ongoing question has been whether AZA treatment clears mutated HSPC clones from the bone marrow facilitating expansion of residual healthy stem cells, or conversely, reprogrammes mutated cells such that blood output is increased. We have previously shown in a smaller MDS/CMML cohort that increased output from mutated progenitors accompanies clinical response ^36^. Here, we tracked mutations in a panel of 111 genes associated with myeloid pathologies (Table S6) across the course of AZA treatment (Table S7). Our cohort had baseline mutational profiles typical of MDS/CMML/AML ^52^ with the most frequent mutations occurring in *TET2* (10/28; 35.7%), *RUNX1* (10/28; 35.7%), *SRSF2* (9/28, 32.1%), and *TP53* (8/28; 28.6%) (Figure S4A, Figure S4B).

Most patients had stable sub-clonal composition across treatment phases irrespective of clinical outcome (Figure 6A). Three patients showed substantially reduced mutational burden corresponding to clinical response at C6D28 with subsequent reappearance of the mutated clone at progression (Figure 6A; blue arrows indicate P20, P18, P06). Conversely, two patients were initially non-responders but showed a delayed response; in both cases mutational burden was initially stable but reduced at the time of response (Figure 6; orange arrows indicate P07, P09). We observed patients with sustained response but fluctuating mutational burden (Figure 6B; P01), patients with stable mutational burden across initial response and subsequent progression (Figure 6B; P05), and patients with decreasing mutational burden in the absence of clinical response (Figure 6B; P08). Overall, mutational burden did not correspond to clinical response, or to global demethylation kinetics (Figure 6B, *lower panels*).

**Figure 6:**
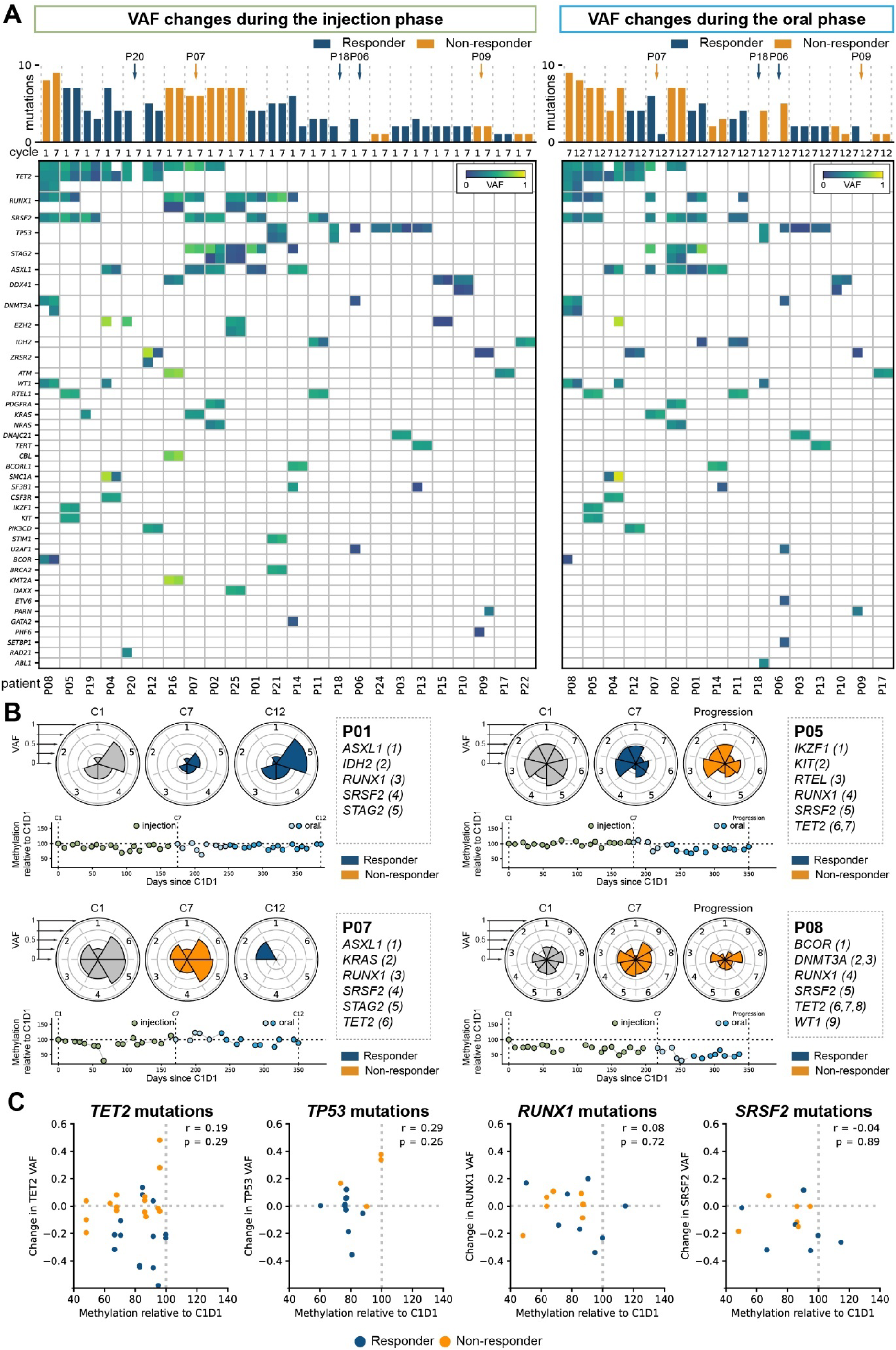
Variations in clonal composition during HMA treatment. **(A)** Variant allele shifts in each patient over (*left*) injection phase, and (*right*) oral phase. Each column shows an individual patient with heatmap representation of VAF for each variant allele at start and end of the treatment phase (or where relevant during the oral phase, at progression). Bar plots are coloured by clinical response across the relevant treatment phase. **(B)** Clonal shifts in representative patients. *Upper panels:* Nightingale plots show VAF for each variant at diagnosis (C1), following 6 treatment cycles (C7) and following 12 treatment cycles/at progression (C12). *Lower panels:* Methylation level relative to C1D1 in PB across the entire course of treatment. **(C)** Correlation between global changes in methylation and shifts in variant allele burden for the four most frequently mutated genes in this cohort. Plots show composite data; each data point indicates a change in VAF across either injection or oral treatment phase and the average global methylation relative to C1D1 over the same treatment phase. r = spearman correlation coefficient.

Finally, we asked whether there was any relationship between shift in specific mutated alleles and global demethylation across the corresponding treatment phase, focussing on the most highly mutated genes. We did not observe any correlation between demethylation and shifts in mutated allele frequency (Figure 6C); furthermore, with the exception of *TET2* (and possibly *TP53),* change in mutation frequency did not differ based on response status (Figure S4C). Thus, although sub-clonal structures of gene mutations showed temporal variation, most variant alleles persisted through both phases of treatment irrespective of clinical response or demethylation kinetics.

## Discussion

In this study we measured longitudinal *in vivo* drug incorporation and DNA methylation dynamics in a cohort of patients treated with injected and oral formulations of AZA. Crossover to oral AZA (100-150mg bd, 21/28days) was generally well tolerated, despite some gastrointestinal toxicities, in a patient cohort with outcomes comparable to historical data^7,8^. Global DNA methylation levels in mononuclear cells were comparable in AZA responders and non-responders during their course of treatment. However, in responders we observed distinct baseline and early drug-induced differences in CpG methylation in CD34^+^ HSPCs that overlapped regulatory regions of genes associated with tissue patterning, cell migration and myeloid differentiation. After six treatment cycles, and concomitant with clinical response, differential hypomethylation in HSPCs mapped to genes involved in positive regulation of hemopoiesis and cell-cell adhesion, suggesting adaptations in the bone marrow as a driver of enhanced haematopoiesis in responders. Overall, these data suggest that specific CpG methylation differences in HSPCs may contribute to variable response to AZA.

Global DNA demethylation in PB and BM MNCs was not proportionate to *in vivo* DAC incorporation, and was not a reliable measure of clinical response. By contrast, HSPCs in responders had lower baseline CpG methylation than in non-responders, and these relatively hypomethylated CpGs mapped to pathways involved in tissue patterning and cell migration. Whilst a larger study is required to ascertain whether baseline CpG methylation at specific sites in HSPCs is predictive of clinical response, these data suggest that HSPCs in some individuals may be epigenetically primed to respond to AZA, and point to pathways that may be associated with such a response. Indeed, hypomethylated DMCs in responders mapped to regions with bivalent histone marks or bound by key hematopoietic TFs in healthy HSPCs ^46^. Genomic regions with these features are associated with driving myeloid and megakaryocyte-erythroid differentiation. The origin of baseline CpG methylation differences in HSPCs, and whether they are related to specific genetic features or the phylogenetic age of the mutated clone is unclear. However, these data provide a foundation for more granular investigations of DNA methylation and variant alleles at single cell resolution ^53^.

In healthy HSPCs, key TFs are pre-emptively bound at regulatory regions of lineage specific genes ^46^. CpG hypomethylation in MDS HSPCs during the first AZA cycle was skewed to non-coding regions, and in responders, mapped to pathways involved in activating myeloid programs. This feature that may have facilitated early myeloid differentiation of these cells. Following multiple cycles of AZA when response was clinically apparent and HSPC populations have likely changed from those at baseline, only a small set of CpGs were hypomethylated, with top enriched pathways including cell-cell adhesion and positive regulation of hemopoiesis and leukocyte differentiation that pointed to altered relationships with other cells in the bone marrow microenvironment.

MDS/CMML are driven by somatic mutations in HSCs, and there is increasing evidence that highly mutated stem cells with altered differentiation capacity contribute to improved peripheral counts in responders ^36^. Indeed, most patients in this cohort had stable clonal composition over treatment phases, regardless of response status, drug incorporation or global demethylation. What remains unclear is whether specific CpG demethylation and improved differentiation capacity occur in most cells, or whether reprogramming of a small fraction of mutant stem cells is sufficient to improve circulating output. This has implications for disease progression as mutated stem cells that do not regain differentiation capacity may persist as a cellular reservoir for progressive disease.

Overall, this study revealed that clinical response was not related to how much global demethylation occurred during treatment, but rather to demethylation of specific CpG sites in HSPCs. Due to limitations with sample material, we only evaluated differences in pooled CD34^+^ fractions using RRBS; genome-wide evaluation of CpG methylation in HSPC subfractions may reveal further insights into differentially methylated CpGs that regulate HMA response. Furthermore, this study focussed on cell-intrinsic effects in HSPCs and their progeny. HMAs potentially affect any replicating cell in the BM, and the interplay between the BM microenvironment and mutated HSPCs are involved in pathogenesis of MDS ^54–57^; thus AZA-induced effects on additional cell types in the bone marrow may modulate clinical response to HMAs. Nevertheless, this study uncovered specific regions in HSPCs where demethylation was associated with therapeutic effects, and such regions are of value not only as a gauge for early HMA response assessment but also as potential sensors for screening novel agents or as targets for site-directed demethylation therapies.

## Online Methods

### Trial design, patients, and oversight

Participants were recruited to an open label phase 2 multicentre investigator-initiated trial (NCT03493646: Evaluating in Vivo AZA Incorporation in Mononuclear Cells Following Vidaza or CC-486). Participants who commenced treatment received 6 cycles of parenteral AZA (75mg/m^2^/day administered subcutaneously for 7 days followed by a rest period of 21 days) followed by 6 cycles of oral AZA (2 cycles at 100mg bid, 4 cycles at 150mg bid for 21 days, followed by a rest period of 7 days) (Figure 1A), and were followed up for a further 12 months. Patients were eligible for enrolment following a new diagnosis of HR-MDS (IPSS; intermediate-2 or high-risk), CMML (bone marrow [BM] blasts 10-29%), or LB-AML (20-30% blasts). 53 patients were assessed for eligibility and 40 commenced treatment (Figure 1B). Pharmacodynamic sampling was performed throughout the treatment period (Figure 1A), and clinical response assessed at the end of cycle 6 (C6D28) and end of cycle 12 (C12D28).

When indicated, best supportive care was allowed in combination with study treatment and included red blood cell or platelet transfusion, antibiotics, antifungals, antivirals, anti-emetics, anti-diarrhoeal agents, and granulocyte colony-stimulating factor (G-CSF). Use of erythropoiesis stimulating agents and hematopoietic growth factors other than G-CSF was excluded. Patients were monitored prior to commencement of each treatment cycle with a physical exam, vital signs, haematology, biochemistry, liver function, and ECOG assessment. Criteria for discontinuation included lack of efficacy, progressive disease, withdrawal by participant, adverse event(s), death, lost follow-up, protocol violation, study termination by the sponsor, pregnancy, recovery, non-compliance with azacitidine, transition to commercially available treatment, physician decision, disease relapse, and symptomatic deterioration. Actual reasons for discontinuation were as shown (Figure 1B).

The trial was funded by Celgene and sponsored by the University of New South Wales. The Kirby Institute, UNSW was responsible for trial coordination. The protocol received ethical approval from the South Eastern Sydney Local Health District Human Research Ethics Committee, and participating sites received Institution approval to conduct the trial prior to commencing recruitment. All participants provided written informed consent. A protocol steering committee oversaw the trial, and independent adjudicators confirmed clinical response assessments provided by attending clinicians. Serious adverse events were recorded during the oral AZA cycles only and were assessed by independent medical monitors. J.O. led the statistical analyses with assistance from J.A.I.T., F.Y., and F.C.K. All authors had access to trial data.

### Primary and secondary trial outcomes

The primary objective was to determine whether there is greater AZA incorporation in DNA following 21 days of oral AZA compared to 7 days of injection AZA in a 28-day treatment cycle, and whether incorporation was associated with greater clinical and/or molecular response.

Secondary objectives of the trial were based on a previous study where patients who responded to injection AZA had a greater fraction of cycling HPCs compared to patients who fail to respond ^16^. However, whether increased replication is associated with increased AZA incorporation was not known. The availability of an assay ^41^ to measure AZA incorporation, and the ability to measure the fraction of replicative HPCs, forms the basis of the secondary objectives of this study.

### Statistical analyses

Statistical comparisons for cell cycle, AZA-MS in BM, LINE-1 qPCR, and VAF data were performed in python (v3.8.3) using the scipy (v1.10.1) stats module. Single timepoint comparisons between outcome groups used the function stats.ttest_ind() while comparisons between timepoints used the function stats.ttest_rel(). Correlations were tested using the function stats.spearmanr(). In all cases, *P* values <0.05 were considered significant.

Drug incorporation into DNA and global DNA methylation levels were compared for responders and non-responders, or for injected and oral phases, using a linear mixed model approach with subject-level random effects to account for within-subject variability. Drug incorporation into PB cells and global DNA methylation levels were compared over collection timepoints (D1, D8, D22) for responders and non-responders, or for injection and oral phases, using a linear mixed model approach with subject-level random effects to account for within-subject variability. Linear mixed model approaches utilised SAS software.

### Processing of BM and PB

Peripheral blood (20mL per timepoint in addition to standard of care collection) was collected in 10mL EDTA vacutainers. Immediately following collection, 1mL of 1mg/mL tetrahydrouridine (THU; abcam #ab142080) was added per tube, and the specimen rocked at 4°C until processing. Blood was centrifuged at 800g for 10min at room temperature, upper plasma layer removed and stored at −80°C, and the remaining red cell/buffy coat fraction treated with two rounds of 1X red blood cell lysis buffer (BD Pharm Lyse #555899). Mononuclear cells (MNCs) were recovered by centrifugation at 800g and pellets snap frozen at −80°C for subsequent DNA and RNA isolation.

Bone marrow aspirates (BMA; 30mL per timepoint in addition to standard of care collection) were collected into 10mL Hanks balanced salt solution (Life Technologies #14170-161) containing 100IU/mL porcine heparin (Pfizer). Immediately following collection, 4mL of 1mg/mL THU was added, and the specimen rocked at 4°C until processing. BMA samples were diluted 1:4 with RPMI medium (Gibco #21870092), underlaid with 10mL Lymphoprep (Axis-Shield #1114547), and centrifuged at 800g for 35min at RT. The upper plasma layer was removed and snap frozen at −80°C, and MNCs collected for storage (pellets for DNA/RNA isolation or viably frozen) or further processing. CD34^+^ cells were isolated using CD34^+^ microbeads (Miltenyi #130-046-703) and an AutoMACS machine (Miltenyi). Cell purities were monitored using flow cytometry (APC-CD34 Miltenyi # 130-113-176) and CD34^+^ and CD34 depleted cells were viably frozen for later use.

### Isolation of DNA and RNA

DNA and RNA were isolated from frozen cell pellets or thawed CD34^+^ cells using either an All-in-One DNA/RNA miniprep kit (BioBasic #BS88203) or an AllPrep DNA/RNA mini kit (Qiagen #80204) and quantified using either a Nanodrop spectrophotometer (Thermo) or a Qubit fluorometer (Thermo) using a Quant iT Qubit dsDNA HS Assay Kit (Invitrogen #Q32851).

### Cell cycle analysis

Up to two million thawed bone marrow mononuclear cells (MNCs) were washed and resuspended in flow cytometry buffer (2% fetal bovine serum, 1mM EDTA in PBS) containing human Fc block (BD Biosciences). Cells were first stained with a PE-Cy5 lineage marker cocktail (CD2 (clone RPA-2.10), CD3 (clone HIT3a), CD10 (clone HI10a), CD19 (clone HIB19), CD20 (cone 2H7) and GPA/CD235ab (clone HIR2); BioLegend) for 15 minutes on ice, then washed and fixed with 1.6% paraformaldehyde for 10 minutes at room temperature and permeabilised using 90% ice-cold methanol for 30 minutes on ice. Fixed, permeabilised cells were then incubated with CD34-PE (clone 8G12, BD Biosciences), CD38-APC/Fire 750 (clone HIT2, BioLegend), and Ki67-AF488 (clone B56, BD Biosciences) antibodies for 1 hour at room temperature in the dark. Cells were washed twice then stained in 0.5µg/mL of 4’,6-diamidine-2-phenylidole dihydrochloride (DAPI, BD Biosciences) for 15 minutes at room temperature and data acquired on a LSRFortessa (BD) flow cytometer at the Mark Wainwright Analytical Centre (UNSW Sydney). Data was analysed using FlowJo software (Tree Star, USA). HSC samples containing less than 50 cells were excluded from analysis.

### Measurement of DAC incorporation and DNA methylation using mass spectrometry (AZA-MS)

DNA DAC incorporation and demethylation was measured by mass spectrometry (AZA-MS) ^41^ at all collection time points. Briefly, 250pmol 5-Aza-2’-deoxy Cytidine-15N4 (15N-DAC; Toronto Research Chemicals # A79695) was spiked-in to 5-10µg purified DNA in a total volume of 30µL. After addition of 10µL of 20mg/mL NaBH_4_, samples were incubated for 20 min at room temperature with agitation. The pH was then neutralised by addition of 1µL 2M HCl. DNA was then fragmented by addition of 30µL of fragmentation mixture (100U/mL benzonase (Sigma-Aldrich #E1014), 240U/mL phosphodiesterase I (Sigma-Aldrich #P3243), 40U/mL alkaline phosphatase (Sigma-Aldrich #P7923), 20mM Tris-HCl pH 8.0, 20mM NaCl, 3mM MgCl_2_) and incubation for one hour at 37°C. Fragmented samples were dried under vacuum (Savant Speedvac Plus SC210A, Thermo) and stored at −80°C prior to mass spectrometry analysis. Standard curves for all measured analytes (Decitabine (DAC; Selleck Chemicals #S1200), Cytidine (C; Selleck Chemicals #S2053), 5-methylcytidine (mC; Cayman Chemical #16111), 2’deoxycytidine (dC; Cayman Chemical #34708), 2’deoxy-5-methylcytidine (mdC; MMP Biomedicals # 0219888310-10mg)) were prepared in parallel and covered the following ranges; DAC, 0.3-40pmol, C/dC, 3-100pmol, mC/mdC, 0.06 – 2pmol, all per 20µL injection.

Samples were reconstituted in 50µL CE buffer (10mM Tris HCl pH 9.0, 0.5mM EDTA) and run on a Q Exactive PLUS Orbitrap mass spectrometry system (Thermo) using a heated electrospray interface operated in the positive ion mode. Chromatographic separation was performed on a 100 mm × 2.1 mm i.d., 3 μM, C30 column (Acclaim, Thermo) at 40 °C. Duplicate 20µL injections of each sample were analysed using gradient elution with 0.1% formic acid in Milli-Q water (Solvent A) and 0.1% formic acid in acetonitrile (Solvent B) at a flow of 0.4 ml/min over 8 min. Mass spectra were acquired at a resolution of 140 000 over the range of 220 to 260 Th with the electrospray voltage at 4000 V. Sheath gas pressure and auxillary gas pressure were 27 and 10. The capillary temperature was 300 °C and the s-lens was 80 V. Data processing of chromatograms was performed using the Quanbrowser function of the Xcalibur Software package version 2.5 (Thermo) and analyte measurements used to calculate pmol DAC detected per µg of input DNA and DNA methylation relative to C1D1 levels ([mdC/dC timepoint] ÷ [mdC/dC C1D1]).

### qPCR-based measurement of DNA demethylation at LINE-1 elements

LINE-1 qPCR was performed essentially as described ^44^ and using the same primers except for a modified HpaII-cut specific reverse F-oligo: TGGCTGTGGGTGGTGGGCCTCGTAGAGGCCCTTTTTTGGTCGGTACCTCAGATGGAAATGTCTT/3ddC/. Technical replicates of up to 4 ng of DNA in PCR master mix (20mM Tris-HCl pH 8.4, 50mM KCl, 7mM MgCl_2_, 0.2mM dNTP mix (dATP, dCTP, dGTP, dTTP), DraI-cut specific oligonucleotides (12.5nM each of forward and reverse F-oligos plus 160nM each of forward and reverse outer primers and HEX probe), HpaII-cut specific oligonucleotides (2nM each of forward and reverse F-oligos plus 40nM each of forward and reverse outer primers and FAM probe), 2U HpaII (NEB #R0171L), 1U DraI (NEB #R0129L), 0.5U Hot Start Taq DNA Polymerase (NEB #M0495L)) were cycled at 37°C for 15 min; 90°C for 5 s; 95°C for 2 min; then 10 cycles of 90°C for 5 s, 95°C for 15 s, 60°C for 1 min, and 68°C for 20 s; followed by 40 cycles of 95°C for 15 s, 65°C for 40 s, and 68°C for 20 s in a CFX96 real-time PCR machine (BioRad). Efficiency of the FAM and HEX reactions was calculated using a standard curve over the range 6.25 pg - 4 ng per reaction, generated from AZA-treated cell-line DNA. The LINE-1 demethylation level for each cycle of each patient was normalized to the corresponding C1D1 sample using the following formula:

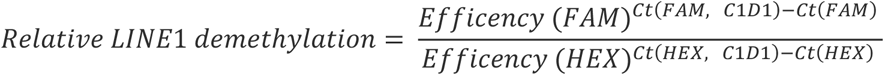

### Reduced representation bisulphite sequencing (RRBS)

RRBS libraries were constructed from 50ng genomic DNA using an Ovation RRBS Methyl-Seq system (Tecan). Bisulphite conversion was performed using the Epitect Fast DNA Bisulfite kit (Qiagen) and libraries amplified using the Ovation RRBS Methyl-Seq system (Tecan). Libraries were sequenced on a Novaseq 6000 using the SP100 flow cell (Illumina) at Monash University’s School of Translational Medicine Genomics Facility.

Raw RRBS data in fastq format were quality and adapter trimmed according to the manufacturer’s guide using trim_galore (0.6.4) and a custom trimming script ^58^. The trimmed fastq files were then aligned to a bisulfite converted genome (hg38) using Bismark (0.22.3) and methylation status at each CpG locus were extracted ^59^. Differentially methylated cytosines (DMCs) were identified using methylKit (1.22.0) incorporating patient ID as a covariate when available and differentially methylated regions (DMRs) were identified with edmr (0.6.4.1) packages in R (4.2.0) ^60,61^.

DMCs and DMRs were annotated using ChIPseeker (1.32.1) for the nearest gene and annotatr (1.22.0) for CpG island annotation ^62,63^. DMCs were further annotated using HiChIP data from healthy HSPC subsets ^46^. DMCs were first linked to significant HiChIP interaction at 5kb resolution, and then associated with genes if the DMC-containing bin was within 10 kb of the TSS of specific genes. Gene ontology enrichment was performed using HiChIP associated genes, and visualized using clusterProfiler (4.11.0.001) ^64^. ChIP-seq peaks ^46^, including TF and histone modifications from different cell types, were used to evaluate the enrichment of DMCs in key regulatory regions. DMCs were first expanded 100bp on both sides and overlapped regions were merged. The regioneR package (1.28.0) was used to generate 100 sets of randomized regions with matched characteristics, and the z-score was calculated as (observe - mean)/sd and visualized ^65^.

### Myeloid capture panel sequencing and analysis

DNA from BM MNCs at baseline and clinical response timepoints was utilised for assessment of VAFs using a myeloid gene capture panel with sensitivity to detect VAFs at or above 5%. Capture sequencing library construction and sequencing was performed at the University of Auckland using standard clinical myeloid capture arrays (Table S7). Sequencing data quality was assessed with fastqc tool (v0.11.5) followed by BWA alignment to the hg19 reference genome (v0.7.12). Sam files were converted to bam format and sorted using samtools (v1.3.1) and mpileup files generated using the following parameters: maximum depth (-d) of 1000, minimum base quality (-Q) of 15 and minimum mapping quality (-q) of 10. Variants were called using Varscan (v 2.3.9) to generate VCF (variant call format) files using the following parameters: minimum coverage (--min) of 8, minimum depth of the reads supporting the variant (--min-reads2) of 2, minimum variant frequency (--min-var-freq) of 0.01 and strand filter (--strand-filter) 0. Variants were annotated using ANNOVAR ^66^ and SnpEffect ^67^.

Variants were filtered to remove those occurring at a minor allele frequency of greater than 0.1% in gnomAD ^68^, ExAC ^69^, and 1000 genomes ^70^ databases. Then any variant belonging to a blacklist of known false positives and any variant at a VAF of less than 5% were excluded.

## Data Availability

All data produced in the present study are available upon reasonable request to the authors

## Acknowledgements

Patients and their carers, clinicians, and trial coordinators at recruiting sites, Celgene/BMS (Dr Jessica Morison, Dr Tamara Etto, Ms Dharini Fernando, Dr Kyle MacBeth, Dr Kevin Lynch, Dr Han Mynt, Dr Du Lam, Ms Tracey Crivellaro, and Dr Amir Samuel), UNSW (Mr Alan Melrose and Dr Qiao Qiao), SESLHD (Ms. Deborah Adrian and Ms Karen Mccardie), and Scientia Clinical Research (Ms Lisa Nelson) for input during protocol development, trial set up and follow-up. Some of the data presented in this work was acquired by personnel and/or instruments at the Mark Wainwright Analytical Centre (MWAC) of UNSW Sydney, which is in part funded by the Research Infrastructure Programme of UNSW. The investigator initiated clinical trial was funded in part by Celgene/BMS (RG172029) with research support from the National Health and Medical Research Council (RG170246, RG211412), Anthony Rothe Memorial Trust (RG182042, RG202657, RG213236), Leukaemia Foundation (RG231257). Patients were eligible for and received injection azacitidine from the pharmaceuticals benefit scheme (Australia) and CC-486 from Celgene/BMS.

## Authorship contributions

J.E.P designed the research study and served as the lead PI on the clinical trial. C.L., wrote the trial protocol and M.N.P. oversaw the clinical trial. J.A.I.T., F.Y. H.R.H, S.J., J.S, C.H.S., X.Y.L., A.C.N., P.M.K, G.S.B., X.Z., M.N., E.S.G., and F.C.K. performed research and analysed data. J.A.I.T., F.V., R.P., M.J.R., C.J.J., S.K.B., D.J.C., J.W.H.W., A.U., M.N.P., and J.E.P. provided research supervision. S.D. and S.H. coordinated the clinical trial. G.B., M.B., Y.G., N.A. provided help with trial coordination and database design. L.V., L.P., C.Y.F., M.K., D.K.H., R.I.S., S.M., L.L., F.H.P., S.R.L., K.J.S., K.K.S., P.R., P.P., W.S.S., S.L., C.T., S.J.F., F.R., A.K.E., D.H. and M.H. assisted with the trial. J.O. performed and oversaw statistical analysis. J.A.I.T and J.E.P. wrote the manuscript. All authors have viewed and approved the manuscript.

