## Supplemental figures for "Clinical Response to Azacitidine in Myelodysplastic Neoplasms is Associated with Distinct DNA Methylation Changes in Haematopoietic Stem and Progenitor cells *in vivo*"

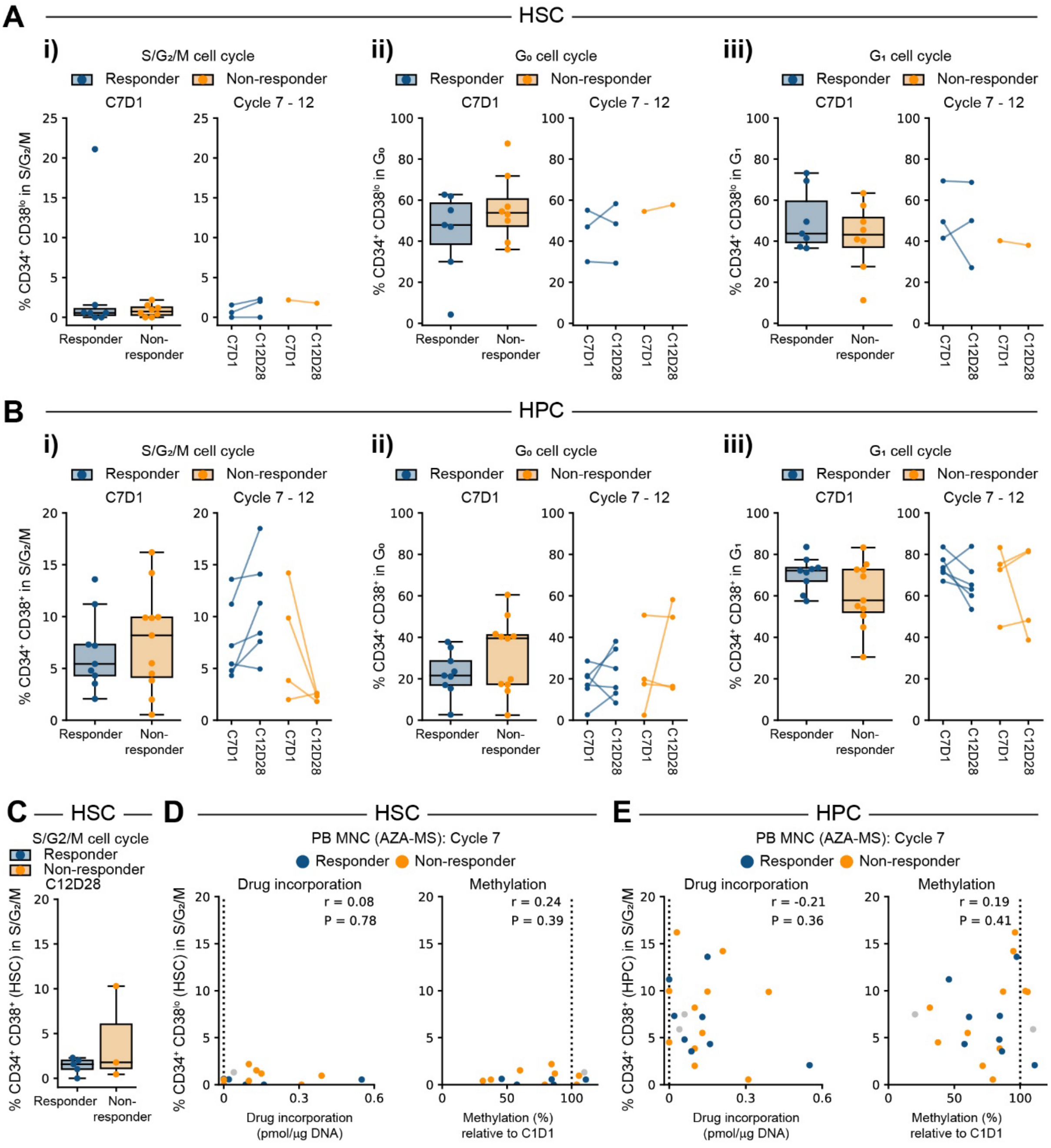
A
HSC
i)
S/G₂/M cell cycle
ii)
G₀ cell cycle
iii)
G₁ cell cycle
Responder
Non-responder
C7D1
Cycle 7 - 12
% CD34⁺ CD38ˡᵒ in S/G₂/M
% CD34⁺ CD38ˡᵒ in G₀
% CD34⁺ CD38ˡᵒ in G₁
C12D28
B
HPC
% CD34⁺ CD38⁺ in S/G₂/M
% CD34⁺ CD38⁺ in G₀
% CD34⁺ CD38⁺ in G₁
C
HSC
S/G2/M cell cycle
% CD34⁺ CD38⁺ (HSC) in S/G₂/M
D
HSC
PB MNC (AZA-MS): Cycle 7
Drug incorporation
Methylation
r = 0.08
P = 0.78
r = 0.24
P = 0.39
% CD34⁺ CD38ˡᵒ (HSC) in S/G₂/M
Drug incorporation
(pmol/µg DNA)
Methylation (%)
relative to C1D1
E
HPC
r = -0.21
P = 0.36
r = 0.19
P = 0.41
% CD34⁺ CD38⁺ (HPC) in S/G₂/M

**Figure S1**

**Figure S1: Cell cycle status at diagnosis and across treatment cycles**

**(A)** Percentage of HSCs in **(i)** $S/G_2/M$, **(ii)** $G_0$, **(iii)** $G_1$ cell cycle phases. **(i-iii)** *Left:* percentage of cells in specified cell cycle phase at C7D1, coloured by clinical outcome at the end of the oral phase. For patients who did not complete 6 cycles, those with progression during the oral phase or soon after stopping treatment were assessed as non-responders (n = 6), and remaining patients were assigned response as per end of cycle 6 IWG assessment (n = 2). *Right:* Percentage of cells in specified cell cycle phase at C7D1 and at the end (C12D28) of the oral phase. Lines indicate paired samples from a single patient, only patients with data at C7 and C12 are shown. *** $P < 0.001$, ** $P < 0.01$, * $P < 0.05$ unpaired (*left*) or paired (*right*) t-test. **(B)** Percentage of HPCs in **(i)** $S/G_2/M$, **(ii)** $G_0$, **(iii)** $G_1$ cell cycle phases. **(i-iii)** *Left:* percentage of cells in specified cell cycle phase at C7D1, coloured by clinical outcome at the end of the oral phase. For patients who did not complete 6 cycles, those with progression during the oral phase or soon after stopping treatment were assessed as non-responders (n = 6), and remaining patients were assigned response as per end of cycle 6 IWG assessment (n = 2). *Right:* Percentage of cells in specified cell cycle phase at C7D1 and at the end (C12D28) of the oral phase. Lines indicate paired samples from a single patient, only patients with data at C7 and C12 are shown. *** $P < 0.001$, ** $P < 0.01$, * $P < 0.05$ unpaired (*left*) or paired (*right*) t-test. **(C)** Percentage of HSCs in $S/G_2/M$ following 12 treatment cycles, coloured by clinical outcome at C12D28. *** $P < 0.001$, ** $P < 0.01$, * $P < 0.05$ unpaired t-test. **(D-E)** Relationship between cell cycle status and drug incorporation and DNA demethylation during cycle 7 in **(D)** HSCs and **(E)** HPCs. **(D-E)** Percentage of cells actively cycling ($S/G_2/M$ phase) at C7D1 compared to maximum drug incorporation (*left*) and minimum relative DNA methylation (*right*) in peripheral blood during the same cycle. Dashed lines indicate baseline values. Gray dots indicate patients for whom response data is unavailable. r = spearman correlation coefficient.

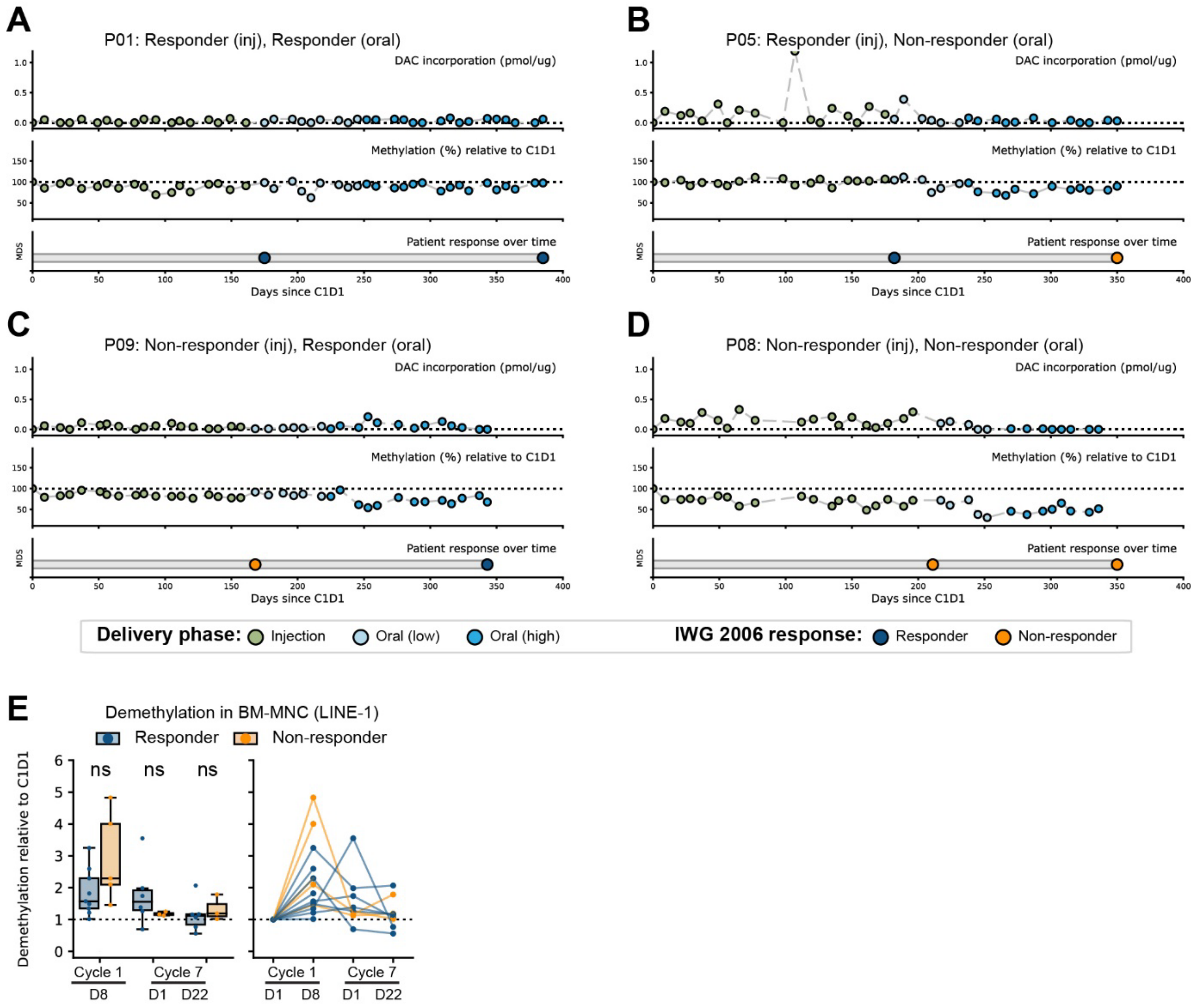
A
P01: Responder (inj), Responder (oral)
DAC incorporation (pmol/ug)
Methylation (%) relative to C1D1
Patient response over time
MDS
Days since C1D1
B
P05: Responder (inj), Non-responder (oral)
DAC incorporation (pmol/ug)
Methylation (%) relative to C1D1
Patient response over time
MDS
Days since C1D1
C
P09: Non-responder (inj), Responder (oral)
DAC incorporation (pmol/ug)
Methylation (%) relative to C1D1
Patient response over time
MDS
Days since C1D1
D
P08: Non-responder (inj), Non-responder (oral)
DAC incorporation (pmol/ug)
Methylation (%) relative to C1D1
Patient response over time
MDS
Days since C1D1
Delivery phase: Injection Oral (low) Oral (high)
IWG 2006 response: Responder Non-responder
E
Demethylation in BM-MNC (LINE-1)
Responder Non-responder
Demethylation relative to C1D1
ns ns ns
Cycle 1 Cycle 7
D8 D1 D22
Cycle 1 Cycle 7
D1 D8 D1 D22

**Figure S2**

**Figure S2: DAC incorporation and relative DNA methylation across treatment phases.** **(A-D)** Full kinetics of DAC incorporation and relative methylation across the complete treatment timeline for 4 participants with differing patterns of clinical response. Circles show response at IWG assessment or progression timepoints (blue – responder, orange – non-responder). **(E)** Relative DNA demethylation in BM-MNC at C1D8, C7D1, and C7D22 compared to pre-treatment. *Left:* aggregate data at each timepoint. *Right:* Plot showing longitudinal methylation changes in individual patients.

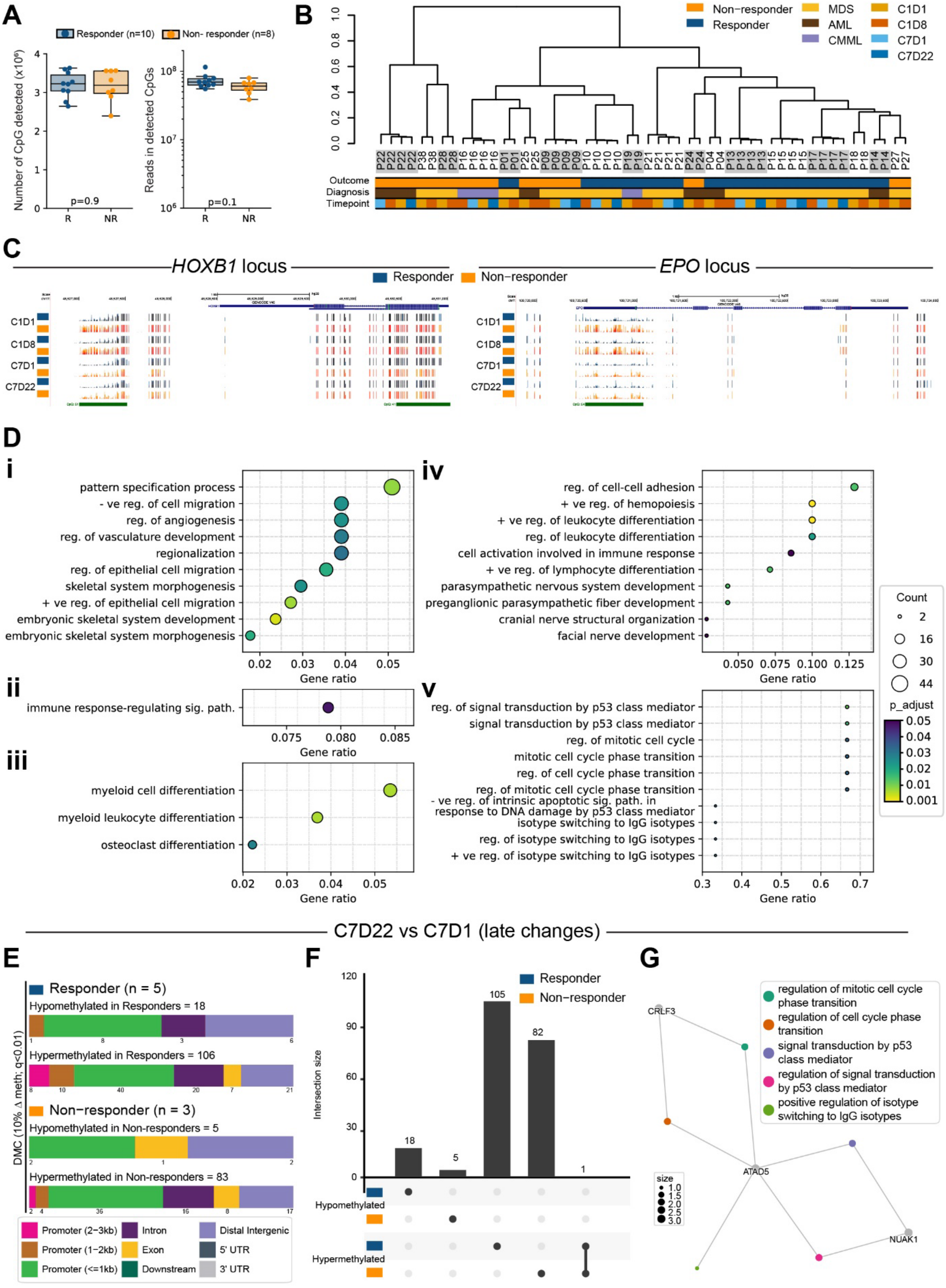
A
Responder (n=10)
Non- responder (n=8)
Number of CpG detected (x10⁶)
Reads in detected CpGs
p=0.9
p=0.1
R
NR
B
Non−responder
Responder
MDS
AML
CMML
C1D1
C1D8
C7D1
C7D22
Outcome
Diagnosis
Timepoint
C
HOXB1 locus
EPO locus
Responder
Non−responder
C1D1
C1D8
C7D1
C7D22
D
i
pattern specification process
- ve reg. of cell migration
reg. of angiogenesis
reg. of vasculature development
regionalization
reg. of epithelial cell migration
skeletal system morphogenesis
+ ve reg. of epithelial cell migration
embryonic skeletal system development
embryonic skeletal system morphogenesis
Gene ratio
ii
immune response-regulating sig. path.
Gene ratio
iii
myeloid cell differentiation
myeloid leukocyte differentiation
osteoclast differentiation
Gene ratio
iv
reg. of cell-cell adhesion
+ ve reg. of hemopoiesis
+ ve reg. of leukocyte differentiation
reg. of leukocyte differentiation
cell activation involved in immune response
+ ve reg. of lymphocyte differentiation
parasympathetic nervous system development
preganglionic parasympathetic fiber development
cranial nerve structural organization
facial nerve development
Gene ratio
v
reg. of signal transduction by p53 class mediator
signal transduction by p53 class mediator
reg. of mitotic cell cycle
mitotic cell cycle phase transition
reg. of cell cycle phase transition
reg. of mitotic cell cycle phase transition
- ve reg. of intrinsic apoptotic sig. path. in response to DNA damage by p53 class mediator
isotype switching to IgG isotypes
reg. of isotype switching to IgG isotypes
+ ve reg. of isotype switching to IgG isotypes
Gene ratio
Count
2
16
30
44
p_adjust
0.05
0.04
0.03
0.02
0.01
0.001
C7D22 vs C7D1 (late changes)
E
Responder (n = 5)
Hypomethylated in Responders = 18
Hypermethylated in Responders = 106
Non−responder (n = 3)
Hypomethylated in Non-responders = 5
Hypermethylated in Non-responders = 83
DMC (10% Δ meth; q<0.01)
Promoter (2−3kb)
Promoter (1−2kb)
Promoter (<=1kb)
Intron
Exon
Downstream
Distal Intergenic
5' UTR
3' UTR
F
Responder
Non−responder
Intersection size
18
5
105
82
1
Hypomethylated
Hypermethylated
G
regulation of mitotic cell cycle phase transition
regulation of cell cycle phase transition
signal transduction by p53 class mediator
regulation of signal transduction by p53 class mediator
positive regulation of isotype switching to IgG isotypes
CRLF3
ATAD5
NUAK1
size

**Figure S3**

**Figure S3: Changes in site-specific methylation over the course of AZA treatment**

**(A-B)** Baseline RRBS data characteristics. **(A)** *Left:* Total number of CpG detected at C1D1, and *Right:* Total number of reads covering detected CpGs at C1D1. **(B)** Hierarchical clustering of all samples based on the most variable CpGs (top 50% based on standard deviation). **(C)** Representative UCSC tracks showing differentially methylated CpGs at the *HOXB1* (chr17:48,526,692-48,531,309 [hg38]) and *EPO* (chr7:100,719,795-100,724,032 [hg38]) loci. Tracks show composite data for each response group at each time point. **(D) (i-v)** Gene ontology analysis for genes associated with differentially methylated CpGs. **(i)** Pathways enriched for genes associated with differentially methylated CpGs hypomethylated in responders compared to non-responders at baseline C1D1. **(ii)** Pathways enriched for genes associated with differentially methylated CpGs hypomethylated in both responders and non-responders at C1D8 vs C1D1. **(iii)** Pathways enriched for genes uniquely associated with differentially methylated CpGs hypomethylated in responders at C1D8 vs C1D1. **(iv)** Pathways enriched for genes associated with differentially methylated CpGs hypomethylated in responders at C7D1 vs C1D1. **(v)** Pathways enriched for genes associated with differentially methylated CpGs hypomethylated in responders at C7D22 vs C7D1. **(E-G)** Sustained changes in site-specific methylation. **(E)** Genomic distribution of CpG sites differentially methylated at C7D22 compared to C7D1 in responders. A total of 1144775 CpG were analysed in responders, and 1240814 CpG were analysed in non-responders. **(F)** Upset plot showing overlap of differentially methylated CpG sites between comparison groups. **(G)** Network diagram showing enriched pathways for genes mapping to CpGs which are hypomethylated in responders at C7D22 compared to C7D1 (n = 18 CpGs). Network diagrams were created using clusterProfiler. CpGs were annotated to genes using HiChIP data from healthy human HSPC subsets (Subramanian et al., 2023).

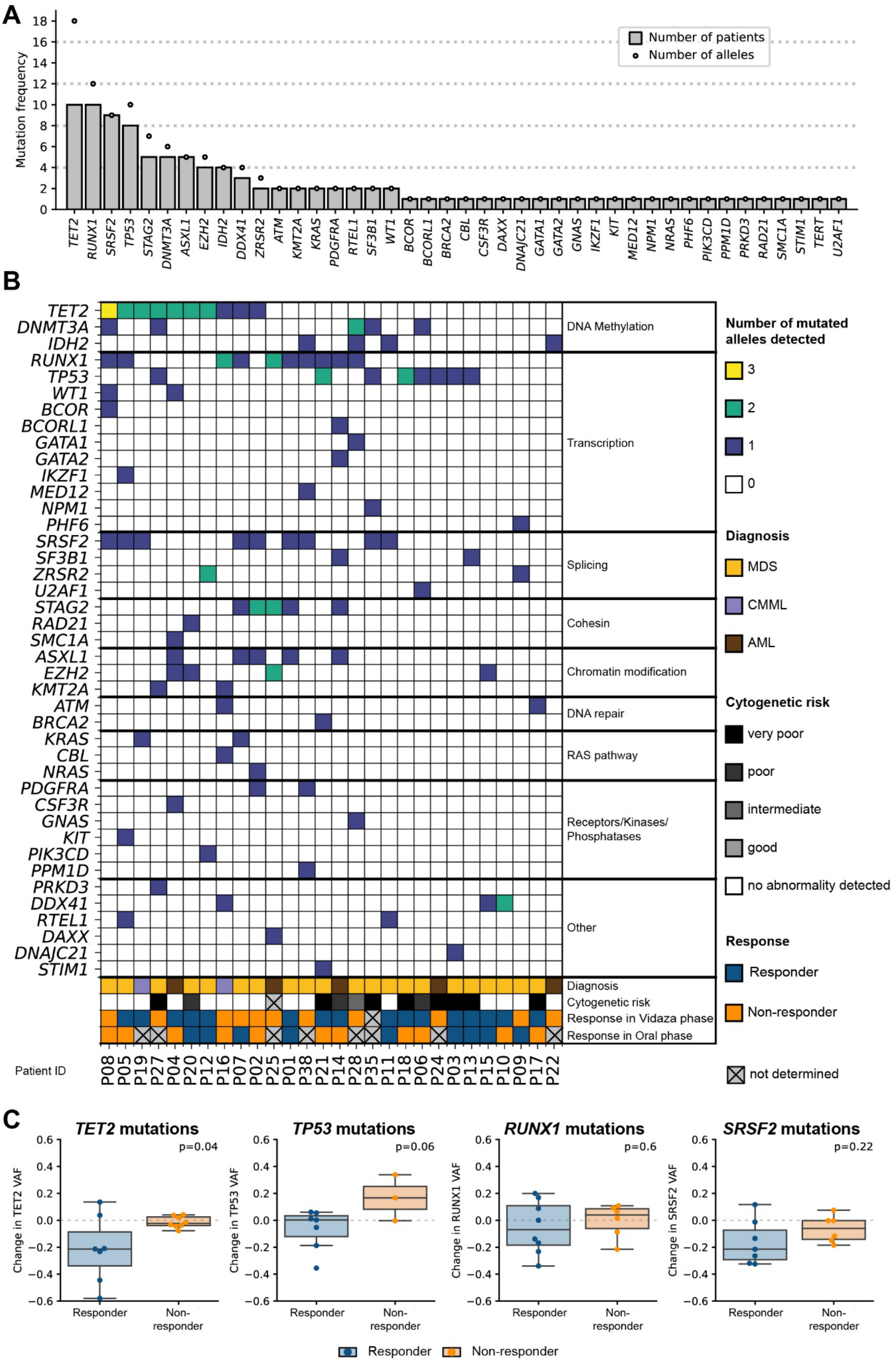
A
Mutation frequency
Number of patients
Number of alleles
TET2 RUNX1 SRSF2 TP53 STAG2 DNMT3A ASXL1 EZH2 IDH2 DDX41 ZRSR2 ATM KMT2A KRAS PDGFRA RTEL1 SF3B1 WT1 BCOR BCORL1 BRCA2 CBL CSF3R DAXX DNAJC21 GATA1 GATA2 GNAS IKZF1 KIT MED12 NPM1 NRAS PHF6 PIK3CD PPM1D PRKD3 RAD21 SMC1A STIM1 TERT U2AF1
B
DNA Methylation
Transcription
Splicing
Cohesin
Chromatin modification
DNA repair
RAS pathway
Receptors/Kinases/ Phosphatases
Other
Diagnosis
Cytogenetic risk
Response in Vidaza phase
Response in Oral phase
Patient ID
P08 P05 P19 P27 P04 P20 P12 P16 P07 P02 P25 P01 P38 P21 P14 P28 P35 P11 P18 P06 P24 P03 P13 P15 P10 P09 P17 P22
Number of mutated alleles detected
3
2
1
0
Diagnosis
MDS
CMML
AML
Cytogenetic risk
very poor
poor
intermediate
good
no abnormality detected
Response
Responder
Non-responder
not determined
C
TET2 mutations
p=0.04
Change in TET2 VAF
TP53 mutations
p=0.06
Change in TP53 VAF
RUNX1 mutations
p=0.6
Change in RUNX1 VAF
SRSF2 mutations
p=0.22
Change in SRSF2 VAF
Responder
Non-responder
Responder
Non-responder

**Figure S4**

**Figure S4: Variations in clonal composition during HMA treatment**

**(A)** Mutation frequency at diagnosis for 28 patients with available data. Grey bars show the number of patients with a mutation detected in the specified gene; open circles indicate the total number of mutated alleles detected in the cohort. **(B)** Overview of mutational profiles in each patient at diagnosis. For each patient, the number of mutated alleles corresponding to each gene is indicated, along with diagnosis, cytogenetic risk score, and response status. Genes are grouped by known function. Cytogenetic risk scores were classified as follows; “no cytogenetic abnormality detected”, “good”: normal karyotype; del(20q); del(5q); del(12p) or double including del(5q), “intermediate”: +8; del(7q); i(17q); +19 or any other single or double independent clone, “poor”: -7; inv(3)/t(3q)/ del(3q); double including -7/del(7q); or complex (3 abnormalities), “very poor”: complex > 3 abnormalities. **(C)** Change in variant allele frequency and patient response status for genes with highest mutation frequency in the cohort. Graphs show combined injection and oral phases, with each data point showing VAF change and corresponding response status. P values shown are for unpaired t-tests.
