## Supplemental tables for "Clinical Response to Azacitidine in Myelodysplastic Neoplasms is Associated with Distinct DNA Methylation Changes in Haematopoietic Stem and Progenitor cells *in vivo*"

**Supplemental tables and figures**

List of Supplemental Tables

Table S1: Baseline demographic and disease characteristics of all participants.

Table S2: Disease response summary

Table S3: IWG2023 classifications

Table S4: Delayed treatment cycles

Table S5: Serious adverse events and adverse events

Table S6: Myeloid gene panel

Table S7: Variant allele frequencies at diagnosis, C6D29, C12D29, and Progression

**Table S1: Baseline demographic and disease characteristics of all participants.** * Cytopenias defined as haemoglobin < 100 g/L, absolute neutrophil count < 1.8 x $10^9$/L, platelet count < 100 x $10^9$/L.

| Characteristic | MDS n= 31 (77.5%) | AML n= 6 (15%) | CMML n= 3 (7.5%) |
|---|---|---|---|
| **Age (yrs) median (min,max)** | 76.2 (65.9, 87.4) | 77.3 (74.9, 83.8) | 73 (73, 82.1) |
| **Gender n(%) Female** | 9 (22.5) | 2 (5) | 1 (2.5) |
| **Male** | 22 (55) | 4 (10) | 2 (5) |
| **ECOG PS n (%)** | | | |
| 0 | 17 (42.5) | 1 (2.5) | 1 (2.5) |
| 1 | 10 (25) | 4 (10) | 2 (5) |
| 2 | 4 (10) | 1 (2.5) | 0 |
| **BMA blast count % median (min,max)** | 12 (1, 20) | 22.5 (20, 30) | 11 (10, 18) |
| **IPSS risk group n= (%)** | | | |
| Low | 0 | | |
| Int-1 | 0 | | |
| Int-2 (1.5-2) | 22 (71) | | |
| High risk (≥2.5) | 9 (29) | | |
| **IPSS-R risk group n= (%)** | | | |
| Very low | 0 | | |
| Low | 1 (3) | | |
| Intermediate | 3 (10) | | |
| High | 15 (48) | | |
| Very high | 12 (39) | | |
| **Karyotype n= (%)** | | | |
| Normal, Y-, 5q-, 20q- | 13 (42) | | |
| Abnormal chromosome 7 or 3 or more abnormalities | 13 (42) | | |
| All other cytogenic abnormalities | 5 (16) | | |
| **Cytopenia of 2 or 3 cell types* n= (%)** | | | |
| No cytopenia or cytopenia of 1 cell type | 6 (19) | | |
| Cytopenia of 2 or 3 cell types | 25 (81) | | |
| **ELN risk stratification n= (%)** | | | |
| Favourable | | 1 (17) | |
| Intermediate | | 3 (50) | |
| Adverse | | 2 (33) | |
| **WHO subtype** | | | |
| CMML-1 blasts | | | 0 (0) |
| CMML-2 blasts | | | 3 (100) |
| **Hematology** | | | |
| Hemoglobin median (min,max) g/L | 96 (73, 132) | 88 (79, 121) | 106 (95, 124) |
| ANC median (min,max) x $10^9$/L | 0.8 (0.2, 26.7) | 1.7 (0.1, 6.7) | 8.9 (4.3, 50.3) |
| WCC median (min,max) x $10^9$/L | 2.3 (0.9, 74.1) | 3.4 (0.7, 23) | 14.8 (8.5, 81.1) |
| Platelet median (min,max) x $10^9$/L | 61 (9, 438) | 43.5 (28, 320) | 40 (23, 99) |

**Table S2: Disease response summary.** Shown are the number of participants achieving the indicated IWG response at end of injected phase (C6) and end of oral phase (C12) and a quantification of best response for each participant who completed the injection phase. Disease progression reported at timepoints other than IWG assessment at C6 and C12 is detailed in Figure 1C.

| | **IWG response** | **C6D29** | **C12D29** | **Best response** |
|---|---|---|---|---|
| Responder | Complete remission (CR) | 7 | 2 | 8 |
| | Partial remission (PR) | 0 | 0 | 0 |
| | Marrow CR | 3 | 4 | 4 |
| | Hematological improvement (HI) | 6 | 1 | 6 |
| Non-responder | Stable disease (SD) | 5 | 2 | 3 |
| | Failure | 1 | 0 | 1 |
| | Relapse after CR or PR | 0 | 1 | 0 |
| | Disease progression | 2 | 1 | 2 |
| Totals | | 24 | 11 | 24 |

**Table S3: IWG2023 classifications.** Compared to IWG2006, classification of one patient (*; P10) changed from non-responder to responder at C12D29.

| Patient | Injection phase | | Oral phase | |
|---|---|---|---|---|
| | **IWG 2023** | **Response** | **IWG 2023** | **Response** |
| P01 | CRbi | Responder | CRh | Responder |
| P02 | No response | Non-responder | | |
| P03 | CR | Responder | CR | Responder |
| P04 | CR | Responder | PD | Non-responder |
| P05 | CR | Responder | Disease relapse | Non-responder |
| P06 | HI-P | Responder | No response | Non-responder |
| P07 | No response | Non-responder | CRuni | Responder |
| P08 | No response | Non-responder | | |
| P09 | No response | Non-responder | CRh | Responder |
| P10 | HI-E | Responder | HI | Responder * |
| P11 | CRbi | Responder | CR | Responder |
| P12 | CR | Responder | CR | Responder |
| P13 | HI | Responder | HI | Responder |
| P14 | CR | Responder | | |
| P15 | HI-P | Responder | | |
| P16 | No response | Non-responder | | |
| P17 | CR | Responder | | |
| P18 | CR | Responder | | |
| P19 | HI | Responder | | |
| P20 | HI-E | Responder | | |
| P21 | CRbi | Responder | | |
| P22 | No response | Non-responder | | |
| P24 | No response | Non-responder | | |
| P25 | No response | Non-responder | | |

**Table S4: Delayed treatment cycles.** Shown are duration and reasons for cycle delays of greater than 3 days.

| Participant | Cycle | Duration (days) | Reason |
|---|---|---|---|
| P01 | 7 | 7 | Participant holidays/work |
| | 8 | 7 | Haematological toxicity |
| | 9 | 14 | Haematological toxicity |
| | 10 | 7 | Haematological toxicity |
| | 11 | 7 | Haematological toxicity |
| | 12 | 7 | Haematological toxicity |
| P02 | 4 | 21 | Non-haematological toxicity |
| | 6 | 28 | SAE/AE followed by suspected disease progression |
| P03 | 7 | 7 | Unable to obtain PD bloods |
| P04 | 1 | 14 | SAE |
| P06 | 2 | 11 | SAE in previous cycle |
| P07 | 9 | 11 | Participant holidays/work |
| P08 | 4 | 28 | Participant holidays/work |
| | 7 | 21 | Haematological toxicity |
| | 10 | 10 | Haematological toxicity |
| P09 | 11 | 7 | SAE |
| P10 | 2 | 7 | Haematological toxicity |
| P14 | 5 | 14 | Xmas closure |
| | 10 | 14 | Suspected disease progression |
| P18 | 7 | 4 | Haematological toxicity |
| P19 | 2 | 7 | Brief hospitalisation for Colitis (Grade 2) |
| P23 | 3 | 21 | Non-haematological toxicity |
| | 5 | 14 | Haematological toxicity |
| P24 | 3 | 7 | SAE |
| | 4 | 8 | Haematological toxicity |
| | 5 | 6 | Chest pain - hospitalisation |
| P32 | 2 | 7 | Non-haematological toxicity |

**Table S5: (A-B) Serious adverse events and adverse events.**

**(A) Serious adverse events (SAE).** Table indicates the number of participants experiencing each SAE in each treatment cycle.

| SAE event | Cycle number - injection phase | | | | | | | Cycle number - oral phase | | | | | | |
|---|---|---|---|---|---|---|---|---|---|---|---|---|---|---|
| | 1 | 2 | 3 | 4 | 5 | 6 | Total | 7 | 8 | 9 | 10 | 11 | 12 | Total |
| febrile neutropaenia | 3 | 3 | 4 | 2 | | | 12 | | | | | 1 | | 1 |
| anaemia | 1 | 1 | 1 | | | | 3 | | | 1 | 1 | | | 2 |
| pyrexia | | | | 1 | | 2 | 3 | 1 | | | | | | 1 |
| thrombocytopenia | 2 | | | 1 | | | 3 | 1 | | | | | | 1 |
| sepsis | | | 1 | | 1 | | 2 | | | | | | | 0 |
| mouth haemorrhage | | 1 | 1 | | | | 2 | | | | | | | 0 |
| musculoskeletal pain | | | 1 | 1 | | | 2 | | | | | | | 0 |
| upper respiratory tract infection | 1 | | 1 | | | | 2 | | | | | | | 0 |
| rectal haemorrhage | | | | 1 | | | 1 | | | 1 | | | 1 | 2 |
| diarrhoea | | | | 1 | | | 1 | | | | 1 | | | 1 |
| neutropenia | | | | 1 | | | 1 | | | | 1 | | | 1 |
| abdominal pain | 1 | | | | | | 1 | | | | | | | 0 |
| acute kidney injury | | | 1 | | | | 1 | | | | | | | 0 |
| acute pulmonary oedema | | | | 1 | | | 1 | | | | | | | 0 |
| anal fissure | 1 | | | | | | 1 | | | | | | | 0 |
| cellulitis | 1 | | | | | | 1 | | | | | | | 0 |
| cerebrovascular accident | 1 | | | | | | 1 | | | | | | | 0 |
| coronary artery disease | | | | 1 | | | 1 | | | | | | | 0 |
| haemolytic anaemia | 1 | | | | | | 1 | | | | | | | 0 |
| haemorrhoid infection | | | 1 | | | | 1 | | | | | | | 0 |
| hepatic failure | | | 1 | | | | 1 | | | | | | | 0 |
| hyponatremia | | 1 | | | | | 1 | | | | | | | 0 |
| intra-abdominal haematoma | 1 | | | | | | 1 | | | | | | | 0 |
| jejunal perforation | | | | 1 | | | 1 | | | | | | | 0 |
| joint swelling | | | 1 | | | | 1 | | | | | | | 0 |
| nasal vestibulitis | | | 1 | | | | 1 | | | | | | | 0 |
| oedema peripheral | 1 | | | | | | 1 | | | | | | | 0 |
| pain | | 1 | | | | | 1 | | | | | | | 0 |
| parotid gland enlargement | | | | 1 | | | 1 | | | | | | | 0 |
| pericardial effusion | | | 1 | | | | 1 | | | | | | | 0 |

| | | | | | | | | | | | | | | |
|---|---|---|---|---|---|---|---|---|---|---|---|---|---|---|
| pneumonia | | | 1 | | | | 1 | | | | | | | 0 |
| septic shock | 1 | | | | | | 1 | | | | | | | 0 |
| skin infection | | 1 | | | | | 1 | | | | | | | 0 |
| soft tissue infection | | | | | 1 | | 1 | | | | | | | 0 |
| transitional cell carcinoma | | 1 | | | | | 1 | | | | | | | 0 |
| upper gastrointestinal haemorrhage | | | | 1 | | | 1 | | | | | | | 0 |
| urinary tract infection | | | | | 1 | | 1 | | | | | | | 0 |
| uterine cancer | | | | | | 1 | 1 | | | | | | | 0 |
| myocardial infarction | | | | | | | 0 | | | 2 | | | | 2 |
| cardiac failure acute | | | | | | | 0 | | | | | | 1 | 1 |
| escherichia sepsis | | | | | | | 0 | | | | 1 | | | 1 |
| lung infection | | | | | | | 0 | | | | | | 1 | 1 |
| prostate infection | | | | | | | 0 | | | 1 | | | | 1 |

**(B) Adverse events: Grade 4 haematological and Grade 3 non-haematological adverse events (AE).** Table indicates the number of participants experiencing each AE in each treatment cycle.

| AE grade | AE event | Cycle number - injection phase | | | | | | | Cycle number - oral phase | | | | | | |
|---|---|---|---|---|---|---|---|---|---|---|---|---|---|---|---|
| | | **1** | **2** | **3** | **4** | **5** | **6** | **Uniq PIDs** | **7** | **8** | **9** | **10** | **11** | **12** | **Uniq PIDs** |
| HAEM AE Grade 4 | neutropenia | 7 | 9 | 8 | 6 | 4 | 4 | 16 | 5 | 3 | 4 | 4 | 3 | 3 | 12 |
| | thrombocytopenia | 11 | 10 | 6 | 4 | 4 | 3 | 15 | 3 | 1 | 2 | 3 | 2 | 2 | 9 |
| | white blood cell count decreased | 1 | 2 | 2 | 2 | 0 | 0 | 3 | 0 | 0 | 0 | 1 | 0 | 1 | 2 |
| Non HAEM AE Grade 3 | abdominal pain | | | | | | | | 0 | 0 | 1 | 0 | 0 | 0 | 1 |
| | anxiety | | | | | | | | 0 | 0 | 1 | 0 | 0 | 0 | 1 |
| | ascites | 0 | 1 | 0 | 0 | 0 | 0 | 1 | | | | | | | |
| | blood creatinine increased | 0 | 0 | 1 | 0 | 0 | 0 | 1 | | | | | | | |
| | chronic obstructive pulmonary disease | 0 | 0 | 0 | 1 | 1 | 0 | 1 | | | | | | | |
| | diarrhoea | | | | | | | | 1 | 1 | 1 | 1 | 0 | 0 | 2 |
| | epistaxis | 0 | 1 | 0 | 0 | 0 | 0 | 1 | | | | | | | |
| | haematuria | | | | | | | | 0 | 1 | 0 | 0 | 0 | 0 | 1 |
| | hyperbilirubinaemia | 0 | 0 | 2 | 0 | 0 | 0 | 2 | | | | | | | |
| | hyperglycaemia | 0 | 0 | 0 | 1 | 0 | 0 | 1 | | | | | | | |
| | hypokalaemia | 0 | 0 | 0 | 1 | 0 | 0 | 1 | | | | | | | |
| | hyponatremia | 0 | 0 | 1 | 0 | 0 | 0 | 1 | | | | | | | |
| | nausea | | | | | | | | 1 | 2 | 0 | 1 | 0 | 0 | 3 |
| | sebaceous carcinoma | 0 | 0 | 0 | 0 | 1 | 0 | 1 | | | | | | | |
| | upper gastrointestinal haemorrhage | 0 | 0 | 0 | 0 | 1 | 0 | 1 | | | | | | | |
| | urinary tract infection | 0 | 1 | 0 | 0 | 0 | 0 | 1 | | | | | | | |
| | vomiting | | | | | | | | 0 | 0 | 0 | 1 | 0 | 0 | 1 |
| | sepsis | | | | | | | | 0 | 0 | 1 | 0 | 0 | 0 | 1 |
| **Total unique PIDs** | | **16** | **19** | **15** | **9** | **8** | **7** | **28** | **8** | **6** | **7** | **7** | **3** | **4** | **19** |

### Table S6: Myeloid gene panel

| Gene Names | | |
|---|---|---|
| *ABCB1* | *GATA1* | *PTEN* |
| *ABCG2* | *GATA2* | *PTPN11* |
| *ABL1* | *GATA3* | *PTPRT* |
| *ALAS2* | *GNAS* | *RAD21* |
| *ANKRD26* | *GNB1* | *RAF1* |
| *ASXL1* | *HIST1H1E* | *RPL11* |
| *ATM* | *HRAS* | *RPL35A* |
| *ATR* | *IDH1* | *RPL5* |
| *AXL* | *IDH2* | *RTEL1* |
| *BCOR* | *IKZF1* | *RUNX1* |
| *BCORL1* | *IL7R* | *SAMD9* |
| *BRAF* | *JAK1* | *SAMD9L* |
| *BRCA1* | *JAK2* | *SAMHD1* |
| *BRCA2* | *JAK3* | *SBDS* |
| *BRD4* | *KDM6A* | *SETBP1* |
| *CALR* | *KIT* | *SF1* |
| *CBL* | *KMT2A* | *SF3A1* |
| *CCND2* | *KRAS* | *SF3B1* |
| *CDA* | *LIG4* | *SMARCA2* |
| *CDKN2A* | *MECOM* | *SMC1A* |
| *CEBPA* | *MED12* | *SMC3* |
| *CSF1R* | *MPL* | *SRP72* |
| *CSF3R* | *MYD88* | *SRSF2* |
| *CTC1* | *MYSM1* | *STAG2* |
| *DAXX* | *NOTCH1* | *STIM1* |
| *DCK* | *NPM1* | *SYK* |
| *DCLK1* | *NRAS* | *TERC* |
| *DDX41* | *PARN* | *TERT* |
| *DIS3* | *PDGFRA* | *TET2* |
| *DKC1* | *PHF6* | *TINF2* |
| *DNAJC21* | *PIK3CD* | *TP53* |
| *DNMT3A* | *PIK3CG* | *TYK2* |
| *ERCC6L2* | *PLCG2* | *U2AF1* |
| *ETV6* | *PPM1D* | *U2AF2* |
| *EZH2* | *PRF1* | *WAC* |
| *FBXW7* | *PRKCB* | *WT1* |
| *FLT3* | *PRKD3* | *ZRSR2* |

**Table S7: Variant allele frequencies at diagnosis, C6D29, C12D29 and Progression**

| Patient ID | Variant | VAF | | | |
|---|---|---|---|---|---|
| | | Diagnosis | C6D29 | Progression | C12D29 |
| P01 | IDH2_p.Arg140Trp | 0 | 0 | | 0.088 |
| | ASXL1_p.Glu635fs | 0.1493 | 0.1232 | | 0.1714 |
| | RUNX1_p.His105Gln | 0.3597 | 0.191 | | 0.3904 |
| | SRSF2_p.Pro95His | 0.3755 | 0.2407 | | 0.3571 |
| | STAG2_p.Ser843fs | 0.6728 | 0.3191 | | 0.7651 |
| P02 | STAG2_p.Ile920fs | 0.0625 | 0.3147 | 0.1698 | |
| | NRAS_p.Gly12Asp | 0.2909 | 0.3776 | 0.3013 | |
| | TET2_p.Arg1235Gln | 0.3822 | 0.3669 | 0.323 | |
| | ASXL1_p.Leu823fs | 0.3952 | 0.4062 | 0.3964 | |
| | PDGFRA_p.Gln722Glu | 0.4703 | 0.3973 | 0.4828 | |
| | SRSF2_p.Pro95Leu | 0.4772 | 0.4746 | 0.3574 | |
| | STAG2_p.Leu609fs | 0.6821 | 0.4233 | 0.3547 | |
| P03 | TP53_p.Pro177His | 0.248 | 0.0606 | | 0.0632 |
| | DNAJC21_p.Lys330Gln | 0.4502 | 0.4566 | | 0.426 |
| P04 | WT1_p.Thr455fs | 0.2944 | 0 | | 0.3361 |
| | TET2_p.Gln1410* | 0.3591 | 0.1382 | | 0.418 |
| | ASXL1_p.Ile395fs | 0.3702 | 0.1975 | | 0.4431 |
| | TET2_p.Gln1701fs | 0.451 | 0 | | 0.4813 |
| | CSF3R_p.His599Asn | 0.4541 | 0.4661 | | 0.5 |
| | EZH2_p.Thr592fs | 0.8202 | 0 | | 0.8577 |
| | SMC1A_p.Arg711Trp | 0.8458 | 0.2163 | | 0.9336 |
| P05 | RUNX1_p.Leu98fs | 0.3684 | 0.136 | | 0.2278 |
| | TET2_p.Arg1237* | 0.4153 | 0.1829 | | 0.2236 |
| | KIT_p.Ala755Val | 0.4615 | 0.4545 | | 0.417 |
| | IKZF1_p.Thr96Met | 0.49 | 0.4377 | | 0.4758 |
| | TET2_. | 0.5123 | 0.304 | | 0.3731 |
| | RTEL1_p.Pro315Arg | 0.5369 | 0.5074 | | 0.5409 |
| | SRSF2_p.Pro95His | 0.5398 | 0.3247 | | 0.322 |
| P06 | ETV6_p.Phe368Leu | 0 | | | 0.1298 |
| | SETBP1_p.Asp868Asn | 0 | | | 0.135 |
| | TP53_p.His179Gln | 0.0532 | | | 0.1667 |
| | U2AF1_p.Gln157Pro | 0.0778 | | | 0.1976 |
| | DNMT3A_p.Arg882Cys | 0.1033 | | | 0.1422 |
| P07 | ASXL1_p.Arg620fs | 0.2607 | 0.2521 | | 0 |
| | KRAS_p.Lys178del | 0.4186 | 0.4173 | | 0.5038 |
| | RUNX1_. | 0.426 | 0.3396 | | 0 |
| | SRSF2_p.Pro95His | 0.4742 | 0.3248 | | 0 |
| | TET2_p.Glu1199fs | 0.6576 | 0.5801 | | 0 |
| | STAG2_p.Arg259* | 0.6867 | 0.6415 | | 0 |

| Patient ID | Variant | VAF | | | |
|---|---|---|---|---|---|
| | | Diagnosis | C6D29 | Progression | C12D29 |
| P08 | DNMT3A_p.Arg13Cys | 0 | 0.2918 | 0.1526 | |
| | RUNX1_p.Arg201Gln | 0.2883 | 0.3956 | 0.1801 | |
| | TET2_p.Lys1329Asn | 0.3036 | 0.3846 | 0.1901 | |
| | BCOR_p.Glu401fs | 0.3051 | 0.0808 | 0 | |
| | SRSF2_p.Pro95Arg | 0.3319 | 0.4062 | 0.2213 | |
| | WT1_p.Ala282Val | 0.3361 | 0.4578 | 0.2073 | |
| | DNMT3A_p.Met548Thr | 0.3577 | 0.4556 | 0.352 | |
| | TET2_p.Gln785fs | 0.3593 | 0.3523 | 0.2521 | |
| | TET2_p.Ser1224Arg | 0.413 | 0.3802 | 0.4177 | |
| P09 | PARN_. | 0 | 0.2692 | | |
| | ZRSR2_p.Ser40* | 0.0644 | 0.064 | | |
| | PHF6_p.Gly10fs | 0.0704 | 0 | | |
| P10 | DDX41_p.Gly548Asp | 0.0669 | 0.0923 | | 0 |
| | DDX41_p.Val463del | 0.2581 | 0.2656 | | 0.2151 |
| P11 | RUNX1_p.Arg427fs | 0 | 0 | | 0.1688 |
| | IDH2_p.Arg140Gln | 0.457 | 0.1958 | | 0.1519 |
| | SRSF2_p.Pro95His | 0.4804 | 0.2155 | | 0.2028 |
| | RTEL1_p.Ala73Ser | 0.5153 | 0.4861 | | 0.5176 |
| P12 | ZRSR2_p.Glu133Lys | 0.2258 | 0 | | 0 |
| | TET2_. | 0.411 | 0.3034 | | 0.3866 |
| | TET2_p.Asn834fs | 0.4206 | 0.207 | | 0.3423 |
| | PIK3CD_p.Glu646Lys | 0.4725 | 0.4236 | | 0.5127 |
| | ZRSR2_p.Glu133fs | 0.8544 | 0.1368 | | 0.2143 |
| P13 | SF3B1_p.Lys700Glu | 0.0657 | 0 | | 0 |
| | TP53_p.Pro151Ser | 0.112 | 0.1667 | | 0.1776 |
| | TERT_p.Ala184Thr | 0.4554 | 0.4466 | | 0.4883 |
| P14 | STAG2_. | 0.0761 | 0 | 0 | |
| | GATA2_p.Gly320Asp | 0.1202 | 0 | 0 | |
| | RUNX1_p.Tyr287* | 0.1392 | 0 | 0 | |
| | SF3B1_p.Lys700Glu | 0.4293 | 0 | 0.1296 | |
| | ASXL1_p.Ala1172Thr | 0.4803 | 0.5 | 0.4808 | |
| | BCORL1_p.Arg1333Lys | 0.5278 | 0.5541 | 0.5129 | |
| P15 | EZH2_p.Arg658Thr | 0.0529 | 0.0513 | | |
| | DDX41_p.Arg543His | 0.1362 | 0.0563 | | |
| P16 | RUNX1_p.His242Arg | 0.0753 | 0.0737 | | |
| | DDX41_p.Glu286Lys | 0.2658 | 0.3307 | | |
| | TET2_p.Gln1653* | 0.4449 | 0.4649 | | |
| | RUNX1_p.His242fs | 0.5127 | 0.5769 | | |
| | CBL_p.Cys381Ser | 0.7123 | 0.7903 | | |
| | ATM_p.Pro1374Leu | 0.7565 | 0.7897 | | |
| | KMT2A_p.Met2783Ile | 0.845 | 0.8007 | | |
| P17 | ATM_p.Pro960His | 0.3538 | 0.3926 | 0.4292 | |

| Patient ID | Variant | VAF | | | |
|---|---|---|---|---|---|
| | | Diagnosis | C6D29 | Progression | C12D29 |
| P18 | ABL1_p.Ser554Cys | 0 | 0 | 0.344 | |
| | WT1_p.Pro136Arg | 0 | 0 | 0.22 | |
| | TP53_p.Cys238Tyr | 0.3562 | 0 | 0.3761 | |
| | TP53_p.Ser106Arg | 0.4032 | 0 | 0.3867 | |
| P19 | TET2_p.Ile1370fs | 0.2973 | 0.0874 | | |
| | KRAS_p.Gly12Arg | 0.3765 | 0 | | |
| | TET2_p.Gly1309Val | 0.4922 | 0.176 | | |
| | SRSF2_p.Pro95His | 0.5424 | 0.2227 | | |
| P20 | RAD21_p.Glu399fs | 0.3525 | | | |
| | TET2_. | 0.4352 | | | |
| | TET2_p.Gln984* | 0.4453 | | | |
| | EZH2_p.Gly628Arg | 0.6437 | | | |
| P21 | TP53_p.Pro301fs | 0.1799 | 0.2402 | | |
| | TP53_p.Arg273His | 0.2479 | 0.3686 | | |
| | STIM1_p.Lys672Arg | 0.5 | 0.5583 | | |
| | BRCA2_p.Lys115Glu | 0.5083 | 0.4818 | | |
| | RUNX1_p.Pro245Leu | 0.5849 | 0.6727 | | |
| P22 | IDH2_p.Arg172Lys | 0.403 | 0.4767 | | |
| P24 | TP53_p.Pro151Ser | 0.2209 | 0.2177 | | |
| P25 | STAG2_p.Phe466fs | 0.1038 | 0.0728 | | |
| | STAG2_p.Ser471Asn | 0.1152 | 0.0876 | | |
| | RUNX1_p.Tyr377* | 0.1602 | 0.1752 | | |
| | EZH2_p.Glu211fs | 0.3367 | 0.4128 | | |
| | RUNX1_. | 0.4115 | 0.4191 | | |
| | EZH2_p.Leu674Ser | 0.4599 | 0.4256 | | |
| | DAXX_p.Pro540Leu | 0.5184 | 0.4898 | | |
| P27 | TET2_p.Ile1918Asn | 0.453 | | | |
| | DNMT3A_. | 0.457 | | | |
| | TET2_p.Thr1916fs | 0.4956 | | | |
| | KMT2A_p.Val2596Ala | 0.5242 | | | |
| | PRKD3_. | 0.6 | | | |
| | TP53_p.Arg273His | 0.9437 | | | |
| P28 | RUNX1_p.Arg166Gln | 0.1286 | | | |
| | DNMT3A_p.Leu723Pro | 0.212 | | | |
| | IDH2_p.Arg172Lys | 0.2816 | | | |
| | DNMT3A_p.Phe734Ile | 0.2886 | | | |
| | GNAS_p.Pro96Ala | 0.4894 | | | |
| | GATA1_p.Gly89Glu | 0.5272 | | | |
| P35 | SRSF2_. | 0.0674 | | | |
| | DNMT3A_p.Lys343fs | 0.3158 | | | |
| | NPM1_p.Pro191Ser | 0.4051 | | | |
| | TP53_. | 0.4313 | | | |

| Patient ID | Variant | VAF | | | |
|---|---|---|---|---|---|
| | | Diagnosis | C6D29 | Progression | C12D29 |
| **P38** | MED12_p.Leu10Gln | 0.0704 | | | |
| | RUNX1_. | 0.2924 | | | |
| | SRSF2_p.Arg94dup | 0.4352 | | | |
| | IDH2_p.Arg140Gln | 0.4684 | | | |
| | PDGFRA_p.Thr157Ala | 0.486 | | | |
| | PPM1D_p.Pro500Arg | 0.508 | | | |
